# Genetic Architecture of Plasma Amyloid and Tau Biomarkers in the Amish

**DOI:** 10.64898/2026.10.15.26365636

**Authors:** Ping Wang, Dana Z. Jian, Yeunjoo E. Song, Audrey Lynn, Kristy Miskimen, Patrice Whitehead, Alex Gulyayev, Michael B. Prough, Daniel A. Dorfsman, Renee A. Laux, Sarada L. Fuzzell, Dawn Miller, Sherri D. Hochstetler, Penelope Miron, Andrew F. Zaman, Larry D. Adams, Laura J. Caywood, Jason E. Clouse, Sharlene D. Herington, Yining Liu, Noel C. Moore, Paula Ogrocki, Alan J. Lerner, Anthony J. Griswold, Michael L. Cuccaro, Jeffery M. Vance, William K. Scott, Margaret A. Pericak-Vance, Jonathan L. Haines

## Abstract

**Background:** Plasma biomarkers offer a minimally invasive alternative for detecting Alzheimer’s disease (AD) pathology, yet the genetic determinants influencing their levels remain largely underexplored.

**Objective:** To identify genetic variants associated with AD-related plasma biomarker levels in a Midwestern Amish cohort.

**Methods:** We analyzed plasma levels of amyloid beta (Aβ42/Aβ40), phosphorylated tau 181 (p-tau181), and the p-tau181/Aβ42 ratio in the Midwestern Amish. Genome-wide association studies (GWAS) were used to examine associations with common genetic variants, while genetic linkage analysis was used to assess the potential impact of rare variants.

**Results:** GWAS found associations between plasma biomarkers and the *APOE* gene. Additionally, novel candidate genes were implicated, including *MME* for Aβ42/Aβ40 ratio, and *VEGFC* and *LRRTM4* for p-tau181. Linkage analysis identified significant signals for p-tau181 on chromosome 1, potentially involving the *CSF1* and *AHCYL1* genes.

**Conclusions:** The novel genetic variants associated with AD pathology implicate metabolic, synaptic, neuronal, and immune pathways. The use of GWAS and linkage analysis sheds light on the genetic architecture of AD-related plasma biomarkers.

## Background

Early and accurate diagnosis of Alzheimer disease (AD) remains a major public health challenge due to the clinical and pathological complexity of the disease, especially in the preclinical stages. Given the heterogeneity of AD and its overlapping clinical presentations with other neurological disorders, misdiagnosis rates remain notably high; up to 12% - 20% of individuals clinically diagnosed with AD fail to meet pathological criteria upon autopsy.^1,2^ Blood based biomarkers are promising in addressing this challenge because they strongly correlate with disease liability and underlying pathological changes.^3^

The primary pathological hallmarks of AD include aggregates of amyloid beta (Aβ) plaques and tau tangles in the brain.^4,5^ While these biomarkers can be accurately measured through neuroimaging or cerebrospinal fluid (CSF) analysis, these techniques are costly, sometimes invasive, and impractical for long-term monitoring of AD progression. With recent advances in high-throughput measurement techniques and immunoassay development, core AD-related biomarkers can now be quantified in blood with high sensitivity.^6^ Plasma Aβ42/Aβ40 and phosphorylated tau at epitope 181 (p-tau181) have been strongly associated with Aβ positivity and tau deposition. Both biomarkers have shown reasonable accuracy in identifying clinically diagnosed AD and related dementia.^7–9^ Additionally, decreased levels of plasma Aβ42/Aβ40 and elevated levels of plasma p-tau181 are associated with the severity of AD^10,11^ and the rate of disease progression.^12,13^ Furthermore, as Aβ42 and p-tau181 provide complementary, yet different aspects of AD pathology, the p-tau181/Aβ42 ratio has emerged as a commonly used composite biomarker. ^9,14,15^ The plasma p-tau181/Aβ42 ratio can distinguish amyloid positive and amyloid negative individuals.^16,17^

Endophenotypes, such as these biomarkers for AD, are likely to have a strong genetic component, but only a few studies have investigated AD-related biomarkers as outcomes. Recent genome-wide association studies (GWAS) of CSF and plasma biomarkers have identified genetic variants associated with known AD risk loci, such as *APOE*, *BACE1*, and *PSEN2* for Aβ,^18–21^ and *MAPT* for tau.^22,23^ However, these studies often face limitations due to small sample sizes, and genetic linkage studies on AD biomarkers remain scarce, particularly in underrepresented populations. The Amish communities offer a unique opportunity to address these gaps. Because certain rare DNA variants may be more common in the Amish population, compared to larger populations, studying this cohort provides greater statistical power for identifying novel genetic variants associated with plasma biomarker levels. Previous studies on founder populations have successfully identified rare and low-frequency variants associated with AD, underscoring the value of these populations for gene discovery.^24–26^ By combining GWAS and genetic linkage approaches, this study aims to advance understanding of AD pathogenesis by identifying genetic determinants of plasma biomarker levels, including plasma Aβ42/Aβ40, p-tau181, and p-tau181/Aβ42 levels.

## 2. Methods

### Study population

The data used and analyzed came from the Collaborative Amish Aging and Memory Project (CAAMP), a prospective multi-site study focused on understanding successful aging, cognitive aging and decline in Midwestern Amish communities located in Ohio (OH) and Indiana (IN). The recruitment and ascertainment process has been detailed in previous publications.^27–29^ In brief, the study team conducted door-to-door interviews to collect demographic and medical history information, blood samples, and cognitive assessments from CAAMP participants. Additionally, using records from the Anabaptist Genealogy Database,^30^ an all-connecting pedigree was created to enable the determination of kinship among participating Amish individuals. Within this study, 3,244 participants had available genotyping data, and 1,403 participants had data available for at least one biomarker of interest.

### Cognitive phenotypes

All participants underwent cognitive screening assessment at the time of enrollment. A clinical adjudication team consisting of neurologists and neuropsychologists used established cutoff scores to assign a consensus cognitive diagnosis.^31^ Individuals were classified into one of four categories: cognitively unimpaired (CU), mild cognitive impairment (MCI), cognitively impaired but not due to Alzheimer’s disease (CINAD), or cognitively impaired due to AD (AD). A small number of participants who required additional follow-up but could not be initially assigned a cognitive diagnosis were not included in the following analyses.

### Plasma biomarkers and quality control

Plasma biomarker measurement and quality control procedures have been described previously.^28,32^ Briefly, plasma samples were aliquoted in in 500 µL volumes and stored at -80°C until analysis. Biomarker concentrations were quantified on the Quanterix™ HD-X platform. Measurements of Aβ40 and Aβ42 were obtained with the Simoa™ Neurology 3-Plex A (N3PA) assay for samples analyzed before the introduction of the Neurology 4-Plex E (N4PE) assay in 2017, after which the N4PE assay was used. Plasma p-tau181 concentrations were quantified using the Simoa™ p-tau181 Advantage V2 assay. All measured biomarker concentrations exceeded the corresponding assay limits of detection.

Quality control (QC) was conducted separately for each biomarker (Supplemental Figure 1). Each sample was linked with corresponding relevant clinical and demographic information. Samples were removed if participants did not have an available consensus cognitive diagnosis or were non-Amish internal controls. When more than one sample was collected from the same participant on a given date, the measurement with the lowest intra-assay coefficient of variation (CV) was selected. Measurements with an intra-assay CV ≥20% were excluded. We reviewed each biomarker distribution using summary statistics and visual inspection. Outliers were identified based on the Median Absolute Deviation (MAD) method, which is more robust than parametric approaches for skewed data.^33^ For each biomarker, only one to three samples were excluded as outliers. After quality filtering, we retained only the most recent biomarker measurement per individual for this study.

### Genotyping, imputation, and quality control

DNA extracted from 30 mL blood samples was genotyped using either the Illumina Expanded Multi-Ethnic Genotyping Array (MEGAEX) with custom content^34^ or the Illumina Global Screening Array (GSA).^35^ The MEGAEX and GSA arrays provided coverage of more than 2 million markers and approximately 660,000 markers respectively. To improve coverage of genetic variation relevant to the Amish, approximately 6,000 additional variants were added as custom content to the GSA, including more than 1,100 novel variants identified from previous studies in the Amish,^27,31,36,37^ as well as other AD-related variants from the Alzheimer’s Disease Sequencing Project.^38^

Supplemental Figure 2 summarizes the workflow used to prepare genotype data for GWAS and linkage analyses in the Amish population. Genotype data was imputed using Trans-Omics for Precision Medicine (TOPMed) reference panel through the Michigan Imputation Server.^39^ MEGAEX and GSA data underwent separate imputation, using GRCh38 for autosomes and hg19 for the X chromosome. After imputation, QC was conducted for each dataset before merging using PLINK 2.0. These QC steps involved the assessment of family relatedness, inconsistencies in reported sex, and batch effects. Single nucleotide polymorphisms (SNPs) with missingness > 5%, a minor allele frequency (MAF) < 0.05, and a Hardy-Weinberg equilibrium (HWE) *p*-value < 1 x 10^-6^ were removed. Additional QC was performed after merging the two datasets, resulting in more than 13 million SNPs across 2,973 individuals that passed QC. Biomarker specific subsets then underwent additional QC before the GWAS and linkage analyses.

### Statistical analysis

Continuous variables with non-normal distributions were compared across groups using the Kruskal-Wallis test, whereas categorical variables were compared using the Chi-square test. To adjust for multiple comparisons (n = 6 tests) in *post-hoc* pairwise comparisons, *p*-values were adjusted for using the Bonferroni correction method.

Prior to genetic analyses, post-QC plasma biomarkers underwent standardization using a rank-based inverse normal transformation. Given the high degree of relatedness among Amish individuals, the GENetic EStimation and Inference in Structured samples (GENESIS) method was applied to account for population structure.^40^ The GENESIS package was used to perform PC-AiR and PC-Relate. PC-AiR conducts principal component analysis (PCA) to infer ancestry while minimizing confounding effects from family structure.^41^ PC-Relate estimates genetic relatedness, including kinship coefficients, identity-by-descent (IBD) sharing probabilities, and inbreeding coefficients.^42^ In this study, we used principal components (PCs) capturing ancestry-related variation to account for population structure, and the optimal number of PCs was determined when cumulative variance exceeded 80%. To address the potential impact of a founder population, we performed linkage disequilibrium (LD) pruning with a threshold of r^2^ < 0.16 to obtain a set of independent genetic variants for linkage analysis. GWAS was then conducted on all autosomal post-QC SNPs, using the GENESIS R/Bioconductor package.^40^ Each SNP was incorporated separately as a genotype dosage (0, 1, or 2) in the association model. General linear models were employed to evaluate genotype-specific differences in mean biomarker levels, adjusting for covariates including study center, age, sex, cognitive diagnosis, biomarker batches, and ancestry-informative PCs. Statistical significance was defined using a genome-wide suggestive threshold of *p*-value ≤ 1 × 10^−5^ and a genome-wide significance threshold of *p*-value ≤ 5 × 10^−8^. Potential genomic inflation was evaluated using quantile-quantile (Q-Q) plots and the genomic inflation factor (λ).

Linkage analysis of plasma biomarkers was conducted using the MERLIN software package (version 1.1.2)^43^ using a variance component-based approach. This method models the covariance among relatives by partitioning phenotypic variance into three sources: additive genetic variance explained by a putative quantitative trait locus (QTL), residual polygenic variance, and environmental variance^44^. For quantitative traits, no explicit Mendelian mode of inheritance (e.g., dominant or recessive) is specified; instead, a maximum likelihood is applied to estimate the additive variance attributable to a QTL at each genomic location. To account for familial correlations and mitigate the influence of pedigree structure, MERLIN incorporates IBD information from pedigree data, which enables the detection of linkage between genetic markers and plasma biomarker variability. Additionally, MERLIN performs a likelihood ratio test to compare the probability of a trait being linked to a specific locus against the null hypothesis or probability that the trait and locus are not linked. This approach is robust for analyzing extended Amish pedigrees while allowing for adjustment of covariates. A LOD score of ≥ 1.86 was used as suggestive of linkage, while a score of ≥ 3.3 was used as strong evidence of linkage.^45^ Notably, MERLIN’s computational constraints limits analyzed pedigrees to fewer than 30 bits.^43,46^ Due to the large family sizes and multi-generational structure of Amish pedigrees, we utilized the PedCut software to subdivide large pedigrees into smaller subunits, thereby reducing the computational burden.^47^ Sub-pedigrees with bit sizes of 21 and 25 were generated for sensitivity analyses.

## Results

Table 1 summarizes the number of individuals and families included in the GWAS and linkage analyses. Across biomarker-specific data subsets, the average age at last exam ranged from 81.0 to 82.1 years, consistent with older populations commonly studied in AD. Females comprise a slightly larger proportion in each dataset, accounting for approximately 60% of the participants. Across the datasets, 25.7% to 26.2% of participants carried at least one A*POE* ε4 allele.

**Table 1.**
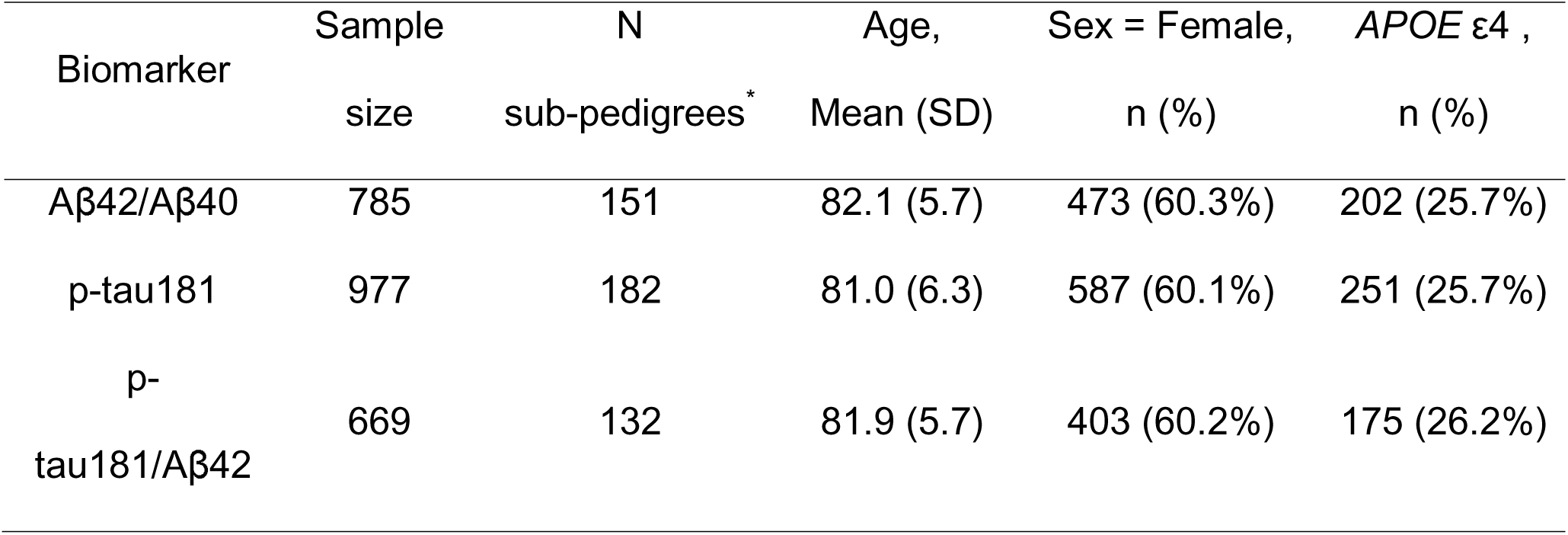
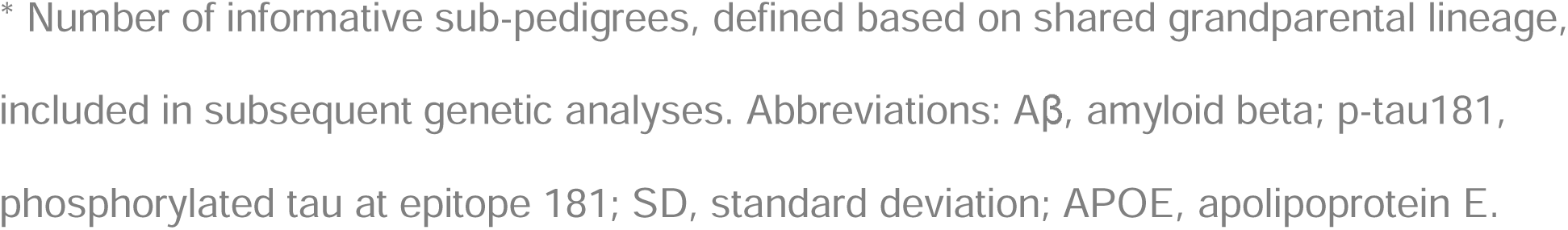
Characteristics of biomarker-specific datasets in the Amish.

| Biomarker | Sample size | N sub-pedigrees* | Age, Mean (SD) | Sex = Female, n (%) | *APOE* ε4 , n (%) |
| --- | --- | --- | --- | --- | --- |
| Aβ42/Aβ40 | 785 | 151 | 82.1 (5.7) | 473 (60.3%) | 202 (25.7%) |
| p-tau181 | 977 | 182 | 81.0 (6.3) | 587 (60.1%) | 251 (25.7%) |
| p-tau181/Aβ42 | 669 | 132 | 81.9 (5.7) | 403 (60.2%) | 175 (26.2%) |

\* Number of informative sub-pedigrees, defined based on shared grandparental lineage, included in subsequent genetic analyses. Abbreviations: Aβ , amyloid beta; p-tau181, phosphorylated tau at epitope 181; SD, standard deviation; APOE, apolipoprotein E.

Table 2 summarizes plasma biomarker levels across cognitive diagnostic groups in the Amish. Although the difference was not statistically significant, the Aβ42/Aβ40 ratio was lower in individuals with AD (0.049 ± 0.012) than in CU individuals (0.052 ± 0.017), consistent with the expected pattern of amyloid pathology in AD. The levels of p-tau181 were significantly higher in the AD group compared with the CU group (2.1 ± 0.9 pg/mL vs 1.9 ± 0.8 pg/mL, *p* = 0.02). The p-tau181/Aβ42 ratio also differed across diagnostic groups, increasing as the severity of AD progressed, with significantly higher levels in the AD group compared with both the CU and CINAD groups after adjustment for multiple comparisons (*p* = 0.002).

**Table 2.**
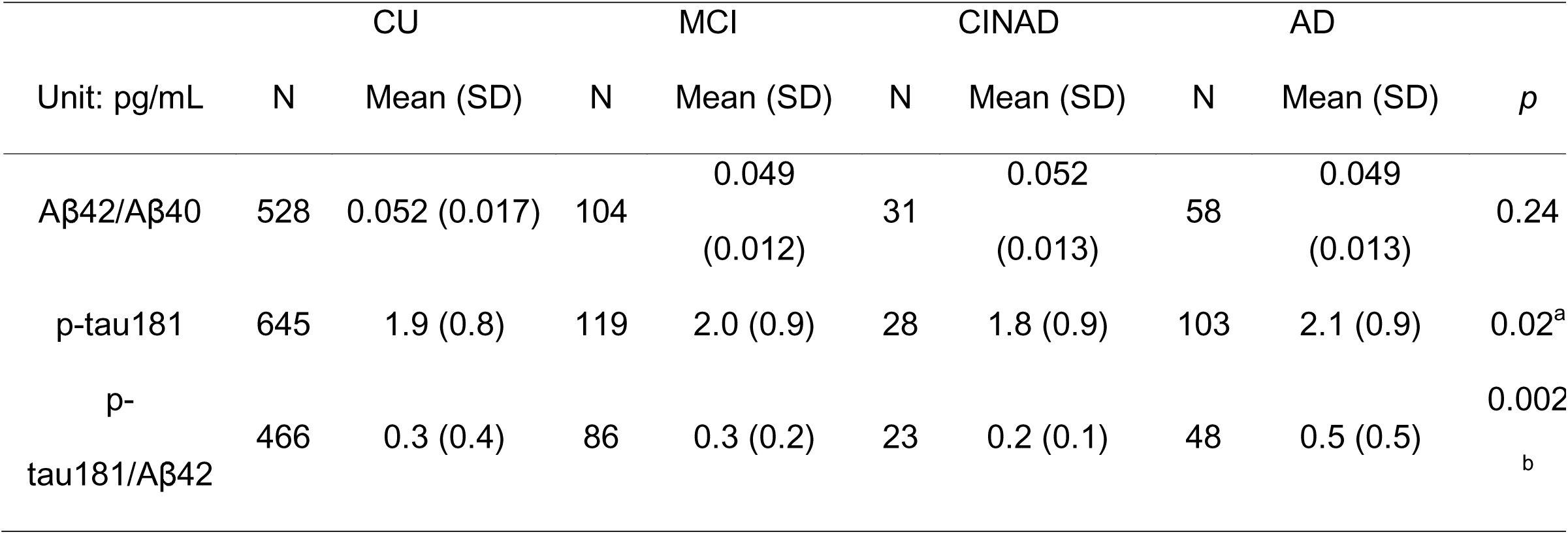
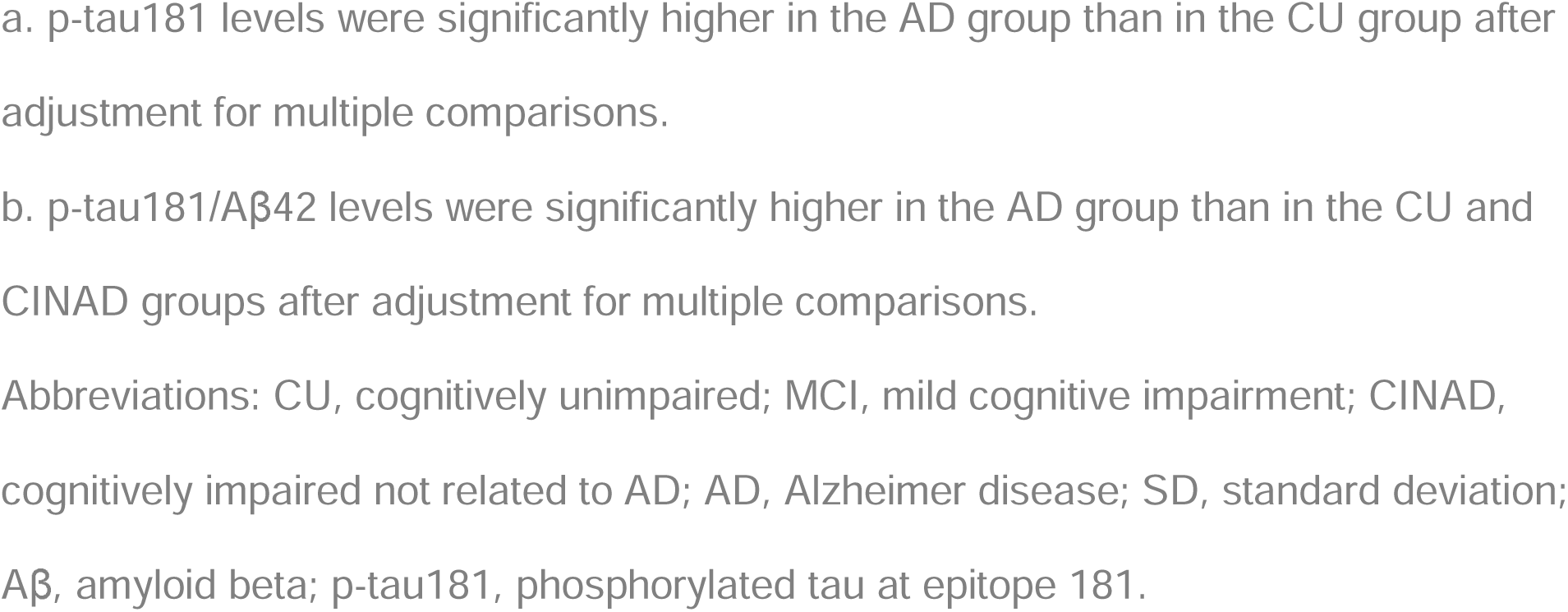
Plasma biomarker levels (pg/mL) across four clinical groups in the Amish.

### Genetic Analyses of Amyloid β Biomarkers

#### Association analysis

The GWAS identified 17 suggestive loci associated with the plasma Aβ42/Aβ40 ratio, with no genomic inflation (λ =1.00) (Figure 1 and Supplemental Table 1). The most significant variant, rs2768733, located on chromosome 10, was associated with an increased plasma Aβ42/Aβ40 ratio (β = 0.29, *p* = 2.07 × 10^⁻7^), along with five additional SNPs demonstrating similar effect sizes. These variants can be mapped to the *FAM107B* (Family with Sequence Similarity 107, Member B) and *ST8SIA6* (ST8 Alpha-N-Acetyl-Neuraminide Alpha-2,8-Sialyltransferase 6) genes. A previously known risk variant within the *APOE* gene, rs429358, demonstrated a suggestive association with a decreased plasma Aβ42/Aβ40 ratio (β = - 0.36, *p* = 2.10 × 10^⁻6^). Furthermore, additional suggestive loci were identified on chromosome 2 near *RAPH1* (Ras Association and Pleckstrin Homology Domains 1; n = 1), chromosome 3 near *MME* (Membrane Metalloendopeptidase; n = 1), and chromosome 4 near *RNU6-1074P* (RNA, U6 Small Nuclear 1074, Pseudogene); n = 8).

**Figure 1.**
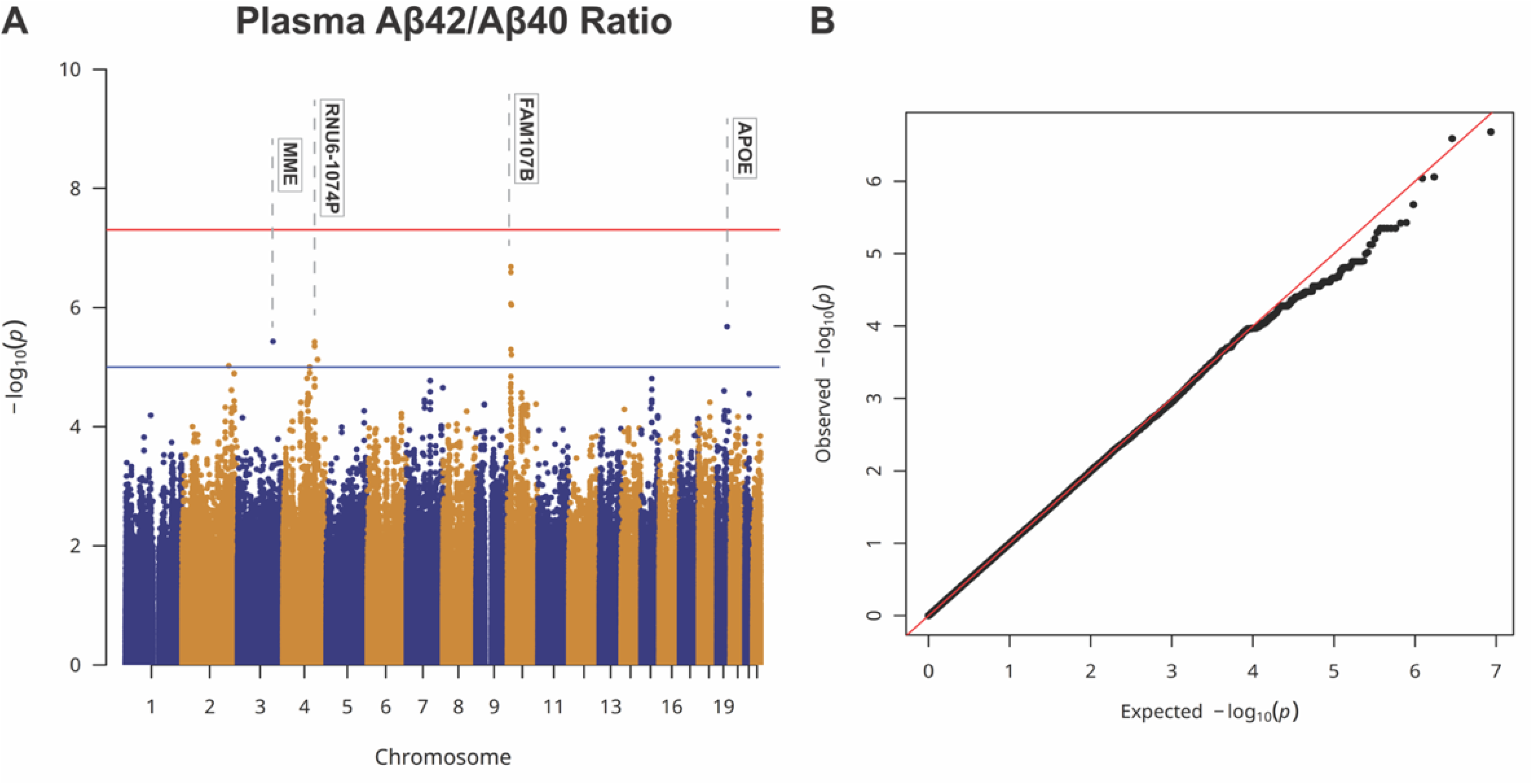
Manhattan plot (A) and quantile-quantile plot (B) of plasma Aβ42/Aβ40 ratio in the Amish. In the Manhattan plot, the blue line marks the genome-wide suggestive threshold (*p* = 1 × 10⁻□), while the red line represents the genome-wide significance threshold (*p* = 5 × 10⁻□). Abbreviations: Aβ, amyloid beta; MME, Membrane Metalloendopeptidase; RNU6-1074P, RNA, U6 Small Nuclear 1074, Pseudogene; FAM107B, Family with Sequence Similarity 107, Member B; APOE, apolipoprotein E.

#### Linkage analysis

Multipoint variance-components linkage analysis of the plasma Aβ42/Aβ40 ratio in 25-bit Amish pedigrees identified suggestive linkage regions on chromosomes 2, 5, and 15 (Figure 2). The strongest linkage signal was identified on chromosome 15 with a peak LOD score of 2.52 at rs4780174, which is located within the *RYR3* (Ryanodine Receptor 3) gene, spanning a 10.8 cM region. Suggestive linkage signals were also observed on chromosome 2, with a peak LOD score of 2.06 at rs199725000 within the long intergenic non-protein coding RNA (*AC006460.1*), and on chromosome 5, with a peak LOD score of 2.26 at rs153512, a variant mapped to another long intergenic non-protein coding RNA (*AC008696.2*). Supplemental Figure 3 presents the linkage results across all autosomes using both 21-bit and 25-bit pedigrees.

**Figure 2.**
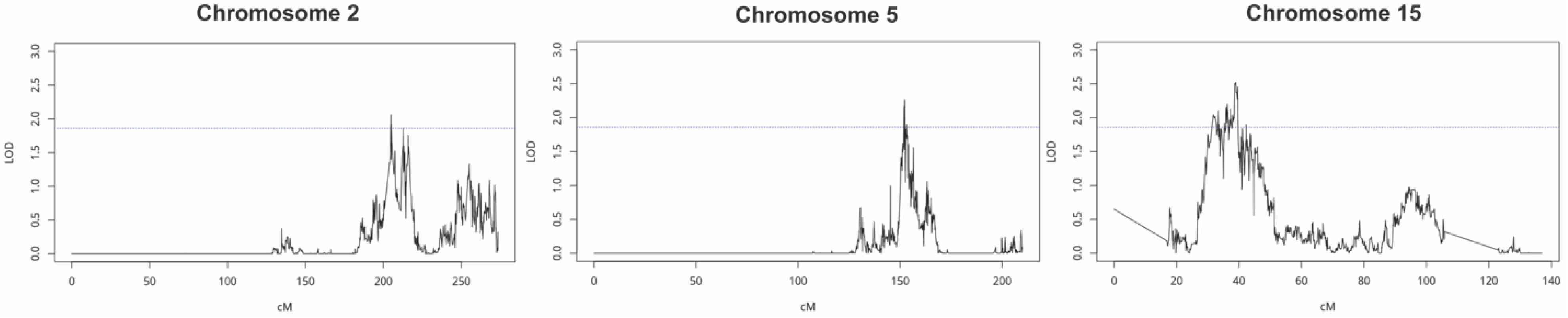
Linkage analysis of plasma Aβ42/Aβ40 ratio using 25-bit pedigrees in the Amish identified 166 suggestive loci (LOD scores ≥ 1.86) on chromosomes 2, 5, and 15. The blue horizontal line represents the suggestive linkage threshold at LOD scores ≥ 1.86. Abbreviations: LOD, logarithm of the odds; cM, centimorgan.

### Genetic analysis of Tau biomarkers

#### Association Analysis

The *APOE* SNP rs429358, associated with the *APOE* ε4 allele on chromosome 19, exhibited the strongest suggestive association with plasma p-tau181 levels in the Amish (β = 0.31, *p* = 1.51 × 10^⁻6^). Additional suggestive loci were found on chromosomes 2 near *MIR4432HG* (MicroRNA 4432 Host Gene) and near *LRRTM4* (Leucine-Rich Repeat Transmembrane Neuronal 4), chromosome 4 near *LTF* (Lactotransferrin), chromosome 6 near *VEGFC* (Vascular Endothelial Growth Factor C), and chromosome 9 near AL161443.1.The GWAS of plasma p-tau181 showed no evidence of genomic inflation (λ = 0.99, Figure 3B). Detailed results for all suggestive loci are provided in Supplemental Table 2.

**Figure 3.**
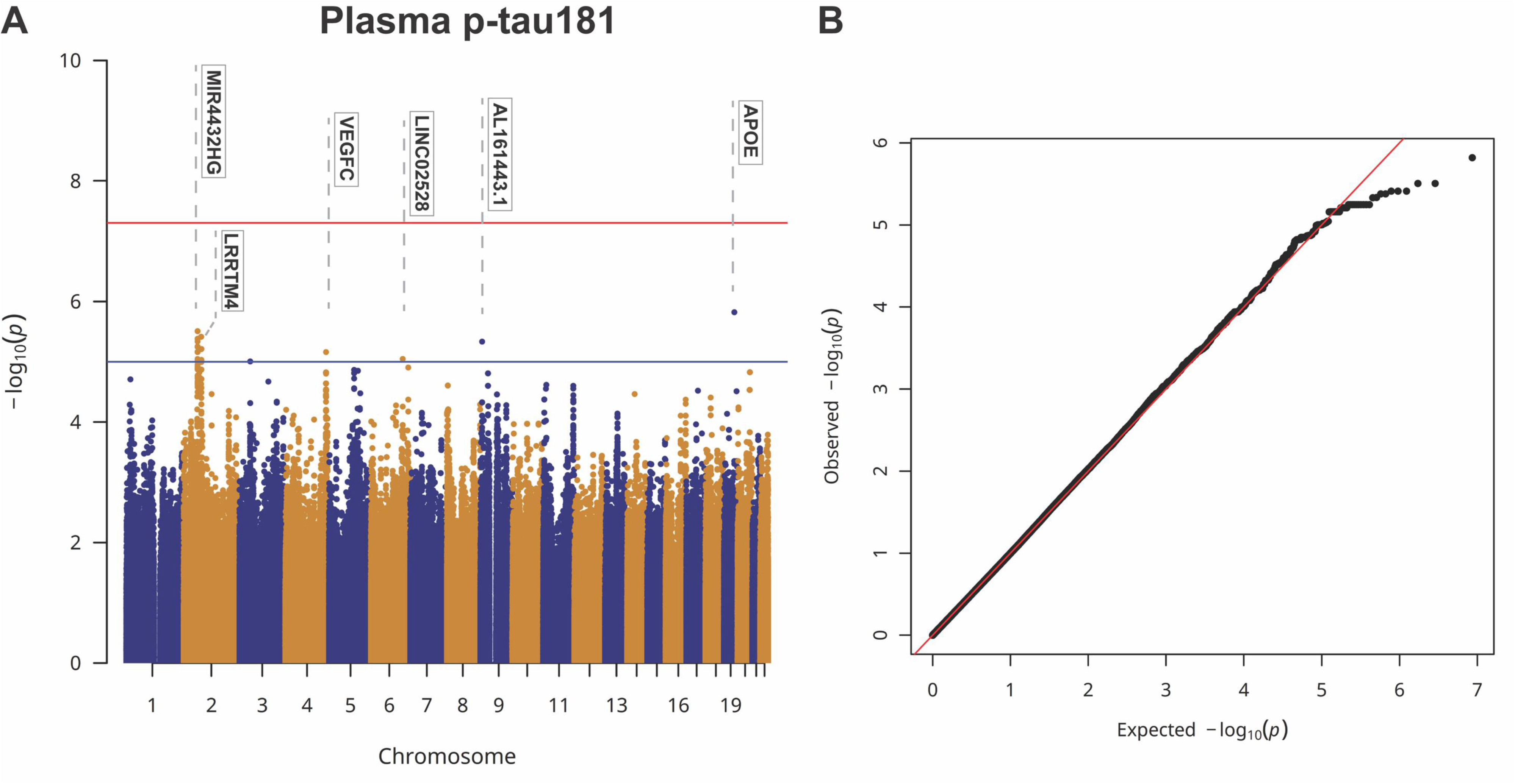
Manhattan plot (A) and quantile-quantile plot (B) of plasma p-tau181 levels in the Amish. In the Manhattan plot, the blue line marks the genome-wide suggestive threshold (*p* = 1 × 10^⁻^), while the red line represents the genome-wide significance threshold (p = 5 × 10^⁻^). Abbreviations: p-tau181, phosphorylated tau at epitope 181; MIR4432HG, MicroRNA 4432 Host Gene; LRRTM4, Leucine-Rich Repeat Transmembrane Neuronal 4; VEGFC, Vascular Endothelial Growth Factor C; LINC02528, Long Intergenic Non-Protein Coding RNA 2528; APOE, apolipoprotein E.

#### Linkage Analysis

Multipoint variance-components linkage analysis of the plasma p-tau181 in 25-bit Amish pedigrees identified a significant linkage region on chromosomes 1 and suggestive linkage regions on chromosomes 9 and 16. In a 56 cM region on chromosome 1 (Figure 4), we identified 16 SNPs showing significant genetic linkage to plasma p-tau181 levels, with the peak signal observed at rs7365459 with a LOD = 3.53. These SNPs are situated within or near several genes, including *CSF1* (Colony Stimulating Factor 1) and *AHCYL1* (S-adenosylhomocysteine Hydrolase Like 1), as well as close to *STRIP1* (Striatin Interacting Protein 1) and *GSTM5* (Glutathione S-Transferase Mu 5). Further suggestive loci were detected on chromosomes 9 and 16 (Supplemental Figure 5). On chromosome 9, the peak LOD score of 2.76 was observed at rs988722022, located within the *TRPM3* (Transient Receptor Potential Cation Channel Subfamily M Member 3) gene. On chromosome 16, the peak LOD score of 2.01 at rs7202587, which maps to the *NPIPB2* (Nuclear Pore Complex Interacting Protein Family Member B2) gene.

**Figure 4.**
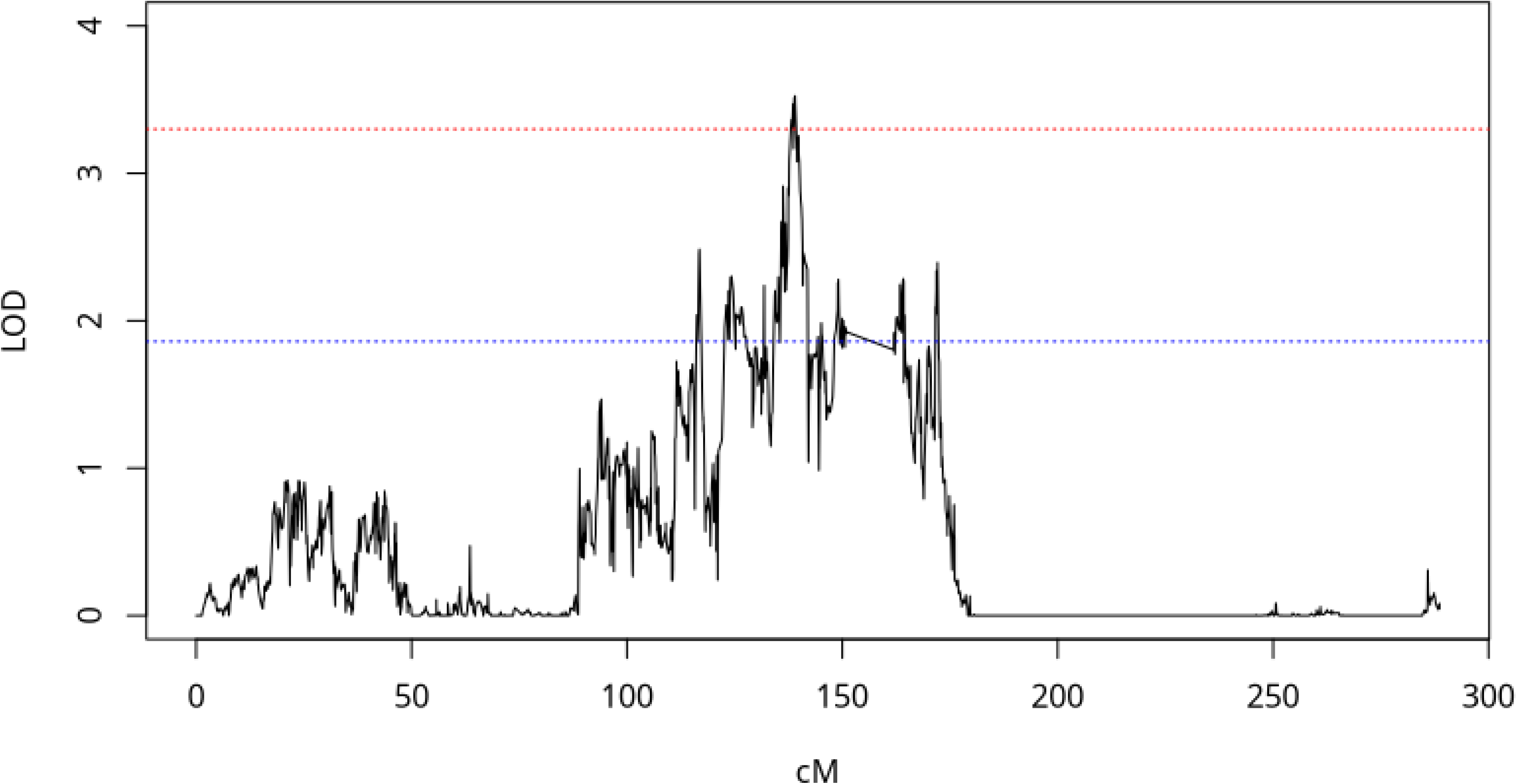
Linkage analysis of plasma p-tau181 levels using 25-bit pedigrees in the Amish identified 16 significant SNPs (LOD scores ≥ 3.3) on chromosome 1. The blue horizontal line represents the suggestive linkage threshold at LOD scores ≥ 1.86, while the red horizontal line marks the significant linkage threshold at LOD scores ≥ 3.3. Abbreviations: LOD, logarithm of the odds; cM, centimorgan.

#### Association and linkage overlap

To integrate the GWAS and linkage findings, association signals within the 1-LOD interval of the chromosome 1 linkage peak for plasma p-tau181 were examined. While none of the linkage loci in this region reached the suggestive threshold in the GWAS, the top association signal for p-tau181, rs61798463 (*p* = 1.25 × 10⁻^□^), lies within the broader linkage peak (approximately 2.9 Mb upstream), indicating overlapping evidence between GWAS and linkage results on chromosome 1 (Supplemental Figure 4).

### Genetic analysis of the composite biomarker

To investigate potential genetic factors that might influence the interplay of Aβ and tau pathologies, we also conducted GWAS and linkage analysis on plasma p-tau181/Aβ42 ratio. No genome-wide significant loci was identified in the GWAS (Supplemental Figure 6). However, the *APOE* locus rs429358 was notably associated with an increased plasma p-tau181/Aβ42 ratio (β = 0.36, *p* = 5.44 × 10^⁻6^). In addition, 26 suggestive loci across chromosomes 4, 6, 8, 9, 11, and 17 were associated with a decreased p-tau181/Aβ42 ratio, with the strongest association observed at rs200118070 on chromosome 4 (β = -0.35, *p* = 1.51 × 10^⁻7^) (Supplemental Table 3). No evidence of genomic inflation was detected (λ = 1.01). Only one locus at rs10033854 showed a suggestive linkage to the plasma p-tau181/ Aβ42 ratio with a LOD score of 1.91 (Supplemental Figure 7). rs10033854 is located near the *UGT2A1* (UDP Glucuronosyltransferase Family 2 Member A1) gene, approximately 48 Mb downstream of the top-associated SNP (rs200118070) identified on chromosome 4 in the GWAS analysis. The complete linkage results for the p-tau181/Aβ42 ratio are provided in Supplemental Figure 8.

Additionally, the association and linkage signals for the p-tau181/Aβ42 ratio was largely distinct, showing minimal overlap with signals obtained from genetic analyses with individual biomarkers (p-tau181 and Aβ42) alone. Although *APOE* SNP rs429358 was suggestively associated with both p-tau181 and the p-tau181/Aβ42 ratio, the remaining suggestive association signals differed between the two biomarkers, as did their linkage patterns. For Aβ42 individually, no genome-wide significant loci was identified in the GWAS (Supplemental Figure 9), although 25 suggestively associated loci across chromosomes 2, 6, 10, 12, 19, 21 were identified. The strongest association was observed at rs72895663 at chromosome 6 (β = 0.26, *p* = 5.14 × 10^⁻7^) (Supplemental Table 4), which is distinct from the chromosome 6 loci identified for the p-tau181/Aβ42 ratio. No evidence of genomic inflation was detected (λ = 0.99) and linkage analysis of Aβ42 alone showed no notable linkage signals.

## Discussion

This study presents a comprehensive genetic analysis of plasma biomarkers associated with AD in the Amish population. Through GWAS and linkage analyses, we identified several suggestive loci linked to the Aβ42/40 ratio and p-tau181. Notably, suggestive associations were found in genes previously implicated in AD pathology, such as *APOE* (rs429358), while novel loci were also potentially implicated. These findings highlight the value of using quantitative traits as outcomes to uncover additional genetic variants influencing AD.

The study also benefits from the unique characteristics of the Amish population, which include both genetic and environmental features. Their relatively homogeneous environment reduces background variation, thereby enhancing the ability to detect genetic contributions to plasma biomarkers. In addition, the founder structure of the Amish population may make it well suited for linkage-based discovery, although the present study did not directly assess whether rare founder variants contribute to the observed linkage signals. To our knowledge, this study is among the first to comprehensively investigate the genetic architecture of blood-based biomarkers in a family-based design, integrating both GWAS and linkage approaches.

The Aβ42/Aβ40 ratio is a stronger indicator of AD pathology than absolute Aβ levels.^7^ In our GWAS involving the Amish population, we found that the *APOE* locus rs429358 SNP was associated with lower ratios, consistent with prior studies indicating a link between *APOE* and increased Aβ aggregation.^18,48,49^ Among the newly implicated genes in this GWAS, rs13081581 within the *MME* gene showed a suggestive association with decreased Aβ42/Aβ40 ratios. *MME* encodes the enzyme neprilysin, an enzyme with an important role in Aβ-degradation in the brain. Reduced neprilysin activity is linked to an increased amyloid burden.^50,51^ Although *MME* was not the primary GWAS hit, its involvement in amyloid clearance warrants further functional studies.

Other genes identified in the analysis include *FAM107B* and *ST8SIA6*; however, neither of them has well-established links to AD or amyloid processing. However, *FAM107B* has been preliminarily linked to AD relevant endophenotypes, including hippocampus and entorhinal cortex volume and oligodendrocyte-associated epigenetic signatures in AD brain tissue.^52,53^ Similarly, *ST8SIA6* may be biologically relevant through its role in sialylation processes that may modulate amyloid aggregation, neuronal or glial function, and its overexpression is associated with anti-inflammatory and neuroprotective effects.^54,55^ Linkage analysis of plasma Aβ42/Aβ40 ratio pointed to *RYR3* on chromosome 15 as a candidate gene. While RYR3 is less studied in the context of AD, its role in calcium homeostasis is crucial for synaptic and neuronal functions.^56^ These results suggest potential mechanisms beyond amyloid precursor processing, although replication studies are needed to confirm the functional role of these novel loci and genes.

Tau pathology is an important component of AD progression, and genetic influences on p-tau levels have been a focus of recent studies. GWAS on CSF and plasma p-tau181 levels have consistently identified *APOE* as the strongest genetic determinant.^18–20,49,57,58^ Additionally, although previous studies have identified associations between *MAPT* and tau, we did not observe a notable *MAPT* signal for plasma p-tau181 in this study. Prior findings suggest that genetic associations with tau may differ across biomarker measures. For example, Chen et al. 2017 found that *MAPT* was associated with plasma total tau, but not with CSF total tau or p-tau.^23^ Similarly, a GWAS of plasma p-tau181 conducted by Lord et al. 2021 identified *APOE*, rather than *MAPT*, as the predominant genome-wide significant signal.^59^ In line with previous findings, GWAS conducted in the Amish also identified the *APOE* association with p-tau181, further reinforcing the well-established role of *APOE* in tauopathy.^60,61^ Notably, we have a suggestive association with p-tau181 and the long non-coding RNA *MIR4432HG.* Although there is currently no strong evidence linking *MIR4432HG* to AD, emerging research on long non-coding RNAs suggests that this could be a promising area for future investigation.^62^ Additionally, suggestive associations near *VEGFC* on chromosome 4 and *LRRTM4* on chromosome 2 indicated that vascular and synaptic factors may contribute to tau pathology. Notably, the suggestive association near *LRRTM4* falls within the chromosome 2 region previously linked to cognitive preservation in the Amish^26^. Specifically, *VEGFC* plays a significant role in brain vascular function, which may influence tau clearance, particularly in the context of AD.^63^ We found SNPs near *LRRTM4* to be negatively associated with p-tau181, suggesting a potential protective effect on tau pathology. *LRRTM4* belongs to the *LRRTM* family of proteins that regulate synaptic functions.^64^ A recent study in the Amish identified a significant genetic linkage locus for cognitive preservation that included *LRRTM4*, with a lead variant predicted to regulate *LRRTM4* expression^26^. Furthermore, another member of this family, *LRRTM3*, has been previously studied for its role in modulating APP processing and Aβ production.^65^ Together, these findings highlight the *LRRTM* family as biologically plausible candidates for protective effects in AD. The linkage analysis identified a significant association between plasma p-tau181 levels and a 56cM region on chromosome 1, with the strongest signal observed at rs7365459 (LOD = 3.53), containing the *CSF1* and *AHCYL1* genes. *CSF1* and its receptor are key regulators of microglial proliferation and function, and they are critical in tau propagation in the brain.^66,67^ While the direct link between *AHCYL1* and AD pathology remains unclear, a recent study demonstrated that *AHCYL1* physically interacts with tau protein, warranting further investigation.^68^ Our findings emphasize the importance of studying neuroinflammation and metabolic regulation as potential contributors to AD pathology.

This study investigated genetic factors influencing the plasma p-tau181/ Aβ42 ratio to better understand the relationship between amyloid and tau pathology in the Amish population. This ratio may improve diagnostic accuracy by accounting for individual variability, potentially providing more consistent indicators of disease state.^9,14,69^

Previous studies on CSF p-tau181/ Aβ42 have revealed strong associations with the *APOE* and *TOMM40* (Outer Mitochondrial Membrane 40) genes.^15,70,71^ We see the *APOE* risk association with an increased plasma p-tau181/Aβ42 ratio in the Amish. Additionally, *NTN1* emerged as a promising candidate gene for this ratio. *NTN1* encodes netrin-1, a protein that directly interacts with APP processing.^72^ It also has neuroprotective properties by mitigating neuronal stress and cell death, making it an intriguing target for AD research.^73,74^ Lastly, linkage analysis of plasma p-tau181/Aβ42 ratio identified a suggestive signal at rs10033854 within the *UGT2A1* gene. Although *UGT2A1* is not recognized as a core AD risk gene, it encodes an enzyme primarily active in the olfactory epithelium.^75^ Given that olfactory impairment is a common early sign in AD, further investigation into UGT2A1 could yield valuable insights into the early mechanisms of AD.^75,76^ Lastly, the plasma p-tau181/Aβ42 ratio showed largely distinct genetic signals compared with p-tau181 and Aβ42 individually, suggesting the ratio may capture genetic influences that are less apparent when its individual components (p-tau181 or Aβ42) is analyzed alone.

Overall, GWAS and linkage findings showed limited overlap across plasma biomarkers, highlighting the distinct aspects of genetic architecture captured by each approach.

GWAS is most effective for identifying common variants, whereas linkage analysis can detect the potential effect of rarer variants segregating in families.^77^ Moreover, large-scale GWAS typically relies on unrelated individuals, whereas linkage analysis considers family structures, potentially capturing long-range haplotype effects not observed in GWAS. The Amish population, with its unique demographic history and reduced genetic diversity, may contain population-specific rare variants that influence biomarker levels. This likely explains why some loci were detected in linkage analyses but not in GWAS, particularly for traits influenced by rare or structural variants that are underrepresented in standard GWAS.

Despite the novelty of this study, several limitations should be acknowledged. Analyzing data within the Amish population may reduce our ability to identify environmental influences on biomarker levels, which could hinder the generalizability of these findings to broader populations. The sample sizes are relatively small for GWAS and linkage analyses, which may reduce the power to detect genome-wide significant signals. While this study identifies several novel suggestive loci for nearly all types of plasma biomarkers, their role in AD pathophysiology remains unclear. Functional validation and replication in independent cohorts will be necessary to clarify the biological implications of these findings. Lastly, it is important to note that plasma biomarkers may not fully reflect underlying central nervous system pathology, and future studies incorporating neuroimaging or cerebrospinal fluid biomarkers may enhance the clinical relevance of these findings.

## Conclusions

In summary, this study presents the most comprehensive genetic investigation of core blood-based AD biomarkers in the Amish population to date. By integrating GWAS and linkage analyses, we demonstrate the complementary strengths of these approaches in identifying genetic factors associated with AD biomarkers. Our findings reveal both novel and previously reported loci that influence Aβ and p-tau181 levels, providing new insights into the mechanisms underlying AD pathophysiology, particularly in pathways related to metabolic regulation, neuronal signaling, synaptic function, and immune responses. Future research should focus on replication in diverse populations and functional validation of candidate loci to translate these genetic discoveries into clinically relevant insights for AD.

## Data Availability

All data produced in the present study may be available upon reasonable request to the authors.

## Acknowledgements

We are deeply grateful to the Amish families for welcoming us into their communities and for their continued participation and contributions to this research.

## Author contributions

### Statements and declarations

#### Ethical considerations

All study procedures were reviewed and approved by the Institutional Review Boards (IRBs) of Case Western Reserve University and the University of Miami and conducted in accordance with institutional and ethical guidelines. Clinical and genetic data were de-identified in accordance with the study protocol to protect participant confidentiality.

#### Consent to participate

Written informed consent was provided by all participants or their legally authorized representative/guardian.

#### Consent for publication

Not applicable.

#### Data availability

The dataset(s) generated and analyzed in this study are not publicly available due to participant privacy and confidentiality considerations. Data may be available upon reasonable request and subject to applicable institutional and study-specific data-sharing requirements.

