## Supplemental Materials for "Genetic Architecture of Plasma Amyloid and Tau Biomarkers in the Amish"

**Supplemental Figure 1.** Biomarker-level quality control

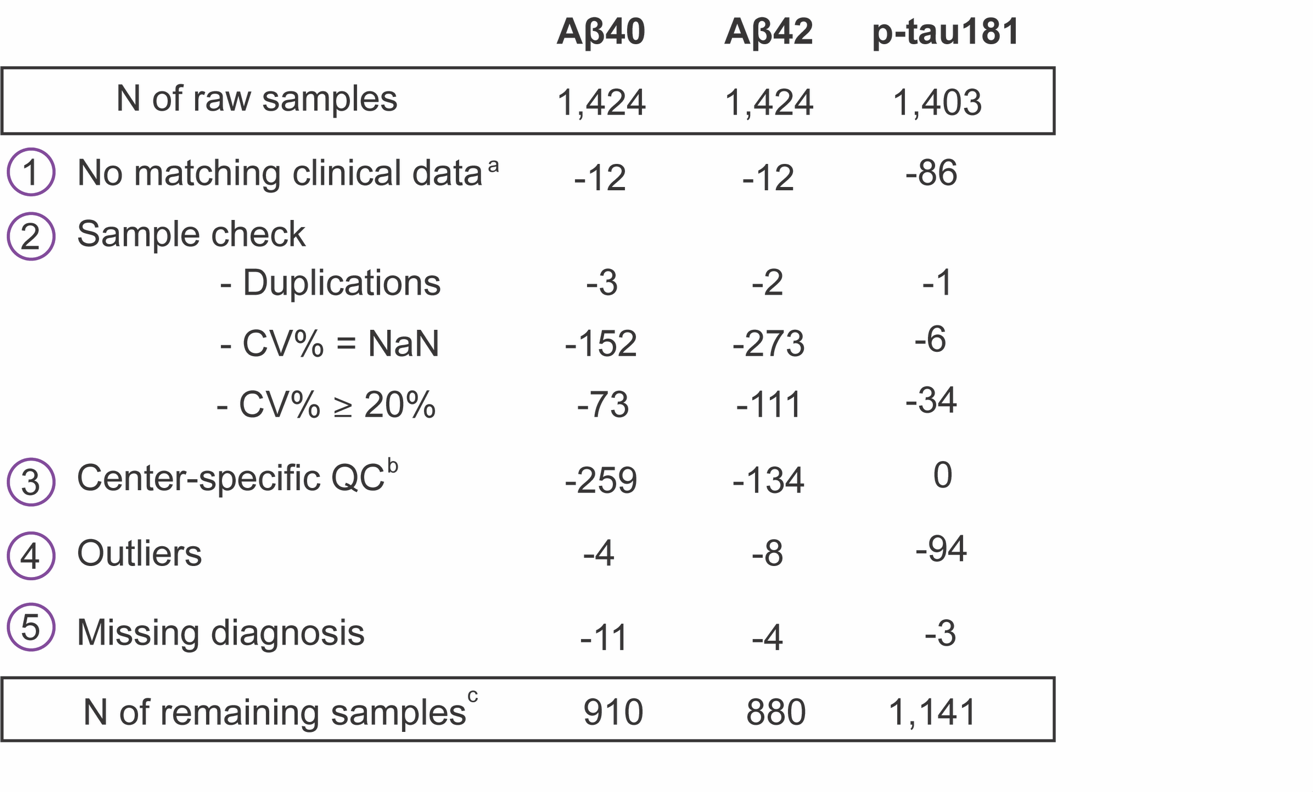

a. Samples had no matching clinical data (i.e., no consensus diagnosis or not Amish).

b. Aβ40 and Aβ42 samples before 2014 from Ohio were excluded.

c. Samples passed QC include one-time and repeated measurements on the same individual.

Abbreviations: CV, Coefficient of variation percentages; QC, quality control; Aβ, amyloid beta; p-tau181, phosphorylated tau at epitope 181.

**Supplemental Figure 2.** Genotyping, imputation, and QC for GWAS and linkage analysis in the Amish.

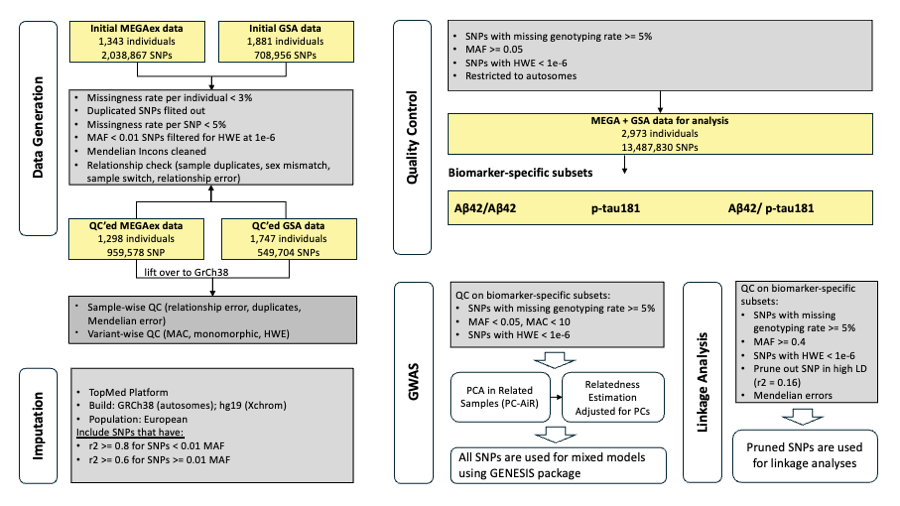

Abbreviations: QC, quality control; GWAS, genome-wide association study; MEGAex, multi-ethnic genotyping array; GSA, global screening array; SNP, single nucleotide polymorphism; MAF, minor allele frequency; MAC, minor allele count; Aβ, amyloid beta; p-tau181, phosphorylated tau at epitope 181; PC, principal components; PCA, principal component analysis; GENESIS, GENetic EStimation and Inference in Structured samples.

**Supplemental Table 1**. SNPs associated with the plasma Aβ42/Aβ40 ratio in the Amish (*p* ≤ 1 × 10⁻⁵).

| **Variant** | **Chr** | **Pos** | **Ref** | **Alt** | **MAF** | **Estimate** | ***p*** | **Overlapped/**  **Nearby Gene** |
| --- | --- | --- | --- | --- | --- | --- | --- | --- |
| rs2768733 | 10 | 14692787 | C | T | 0.28 | 0.293 | 2.07E-07 | FAM107B |
| rs2266374 | 10 | 14694231 | T | C | 0.28 | 0.290 | 2.57E-07 | FAM107B |
| rs2609837 | 10 | 14692493 | C | T | 0.30 | 0.267 | 8.70E-07 | FAM107B |
| rs423631 | 10 | 17304545 | G | A | 0.47 | 0.258 | 9.10E-07 | ST8SIA6 |
| rs429358 | 19 | 44908684 | T | C | 0.14 | -0.359 | 2.10E-06 | APOE |
| rs13081581 | 3 | 155190492 | C | A | 0.45 | -0.237 | 3.72E-06 | MME |
| rs6841480 | 4 | 139246240 | C | A | 0.33 | -0.245 | 3.78E-06 | RNU6-1074P |
| rs13110095 | 4 | 139249126 | A | G | 0.33 | -0.244 | 4.47E-06 | RNU6-1074P |
| rs7669972 | 4 | 139249806 | C | T | 0.33 | -0.244 | 4.47E-06 | RNU6-1074P |
| rs10030899 | 4 | 139250420 | C | A | 0.33 | -0.244 | 4.47E-06 | RNU6-1074P |
| rs10031549 | 4 | 139251928 | A | G | 0.33 | -0.244 | 4.47E-06 | RNU6-1074P |
| rs7694975 | 4 | 139253953 | G | A | 0.33 | -0.244 | 4.47E-06 | RNU6-1074P |
| rs12767375 | 10 | 14683089 | T | C | 0.34 | 0.238 | 5.07E-06 | FAM107B |
| rs405337 | 10 | 17305361 | C | G | 0.44 | 0.233 | 6.24E-06 | ST8SIA6 |
| rs17028449 | 4 | 152035428 | T | G | 0.11 | -0.343 | 7.51E-06 | LINC02273 |
| rs28868301 | 4 | 152043242 | C | T | 0.11 | -0.343 | 7.51E-06 | LINC02273 |
| rs13016495 | 2 | 203581104 | C | T | 0.12 | -0.341 | 9.55E-06 | RAPH1 |

Abbreviations: SNP, single nucleotide polymorphism; Aβ, amyloid beta; Chr, chromosome; Pos, position; Ref, reference allele; Alt, alternative allele; **FAM107B**, Family with Sequence Similarity 107, Member B; **ST8SIA6**, ST8 Alpha-N-Acetyl-Neuraminide Alpha-2,8-Sialyltransferase 6; APOE, apolipoprotein E; **MME**, Membrane Metalloendopeptidase; **RNU6-1074P**, RNA, U6 Small Nuclear 1074, Pseudogene; **RAPH1**, Ras Association and Pleckstrin Homology Domains 1.

**Supplemental Figure 3.** Linkage analysis of the plasma Aβ42/Aβ40 ratio in the Amish using 21-bit (blue) and 25-bit pedigrees (red).

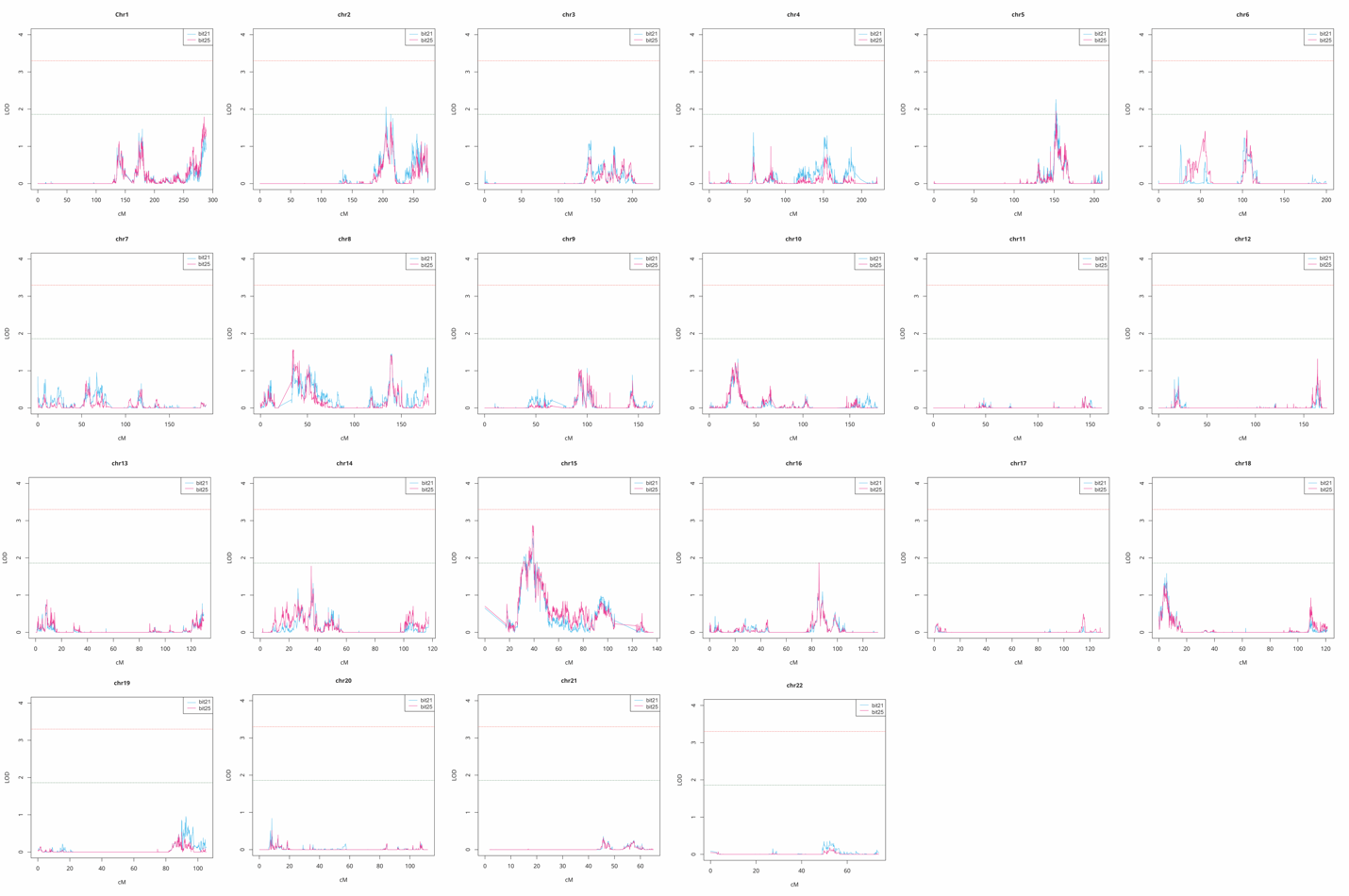

Note: The blue horizontal line represents the suggestive linkage threshold at LOD scores ≥ 1.86, while the red horizontal line marks the significant linkage threshold at LOD scores ≥ 3.3. Abbreviations: Aβ, amyloid beta; LOD, logarithm of the odds; cM, centimorgan.

**Supplemental Table 2**. SNPs associated with plasma p-tau181 levels in the Amish (*p* ≤ 1 × 10⁻⁵).

| **Variant** | **Chr** | **Pos** | **Ref** | **Alt** | **MAF** | **Estimate** | ***p*** | **Overlapped/**  **Nearby Gene** |
| --- | --- | --- | --- | --- | --- | --- | --- | --- |
| rs429358 | 19 | 44908684 | T | C | 0.14 | 0.312 | 1.51E-06 | APOE |
| rs12991515 | 2 | 60421645 | G | A | 0.40 | -0.216 | 3.12E-06 | MIR4432HG |
| rs12997266 | 2 | 60422008 | G | A | 0.40 | -0.216 | 3.12E-06 | MIR4432HG |
| rs1468429 | 2 | 75753500 | A | T | 0.14 | -0.289 | 3.87E-06 | AC005034.4 |
| rs13421024 | 2 | 75768217 | G | C | 0.14 | -0.289 | 3.87E-06 | AC005034.4 |
| rs10172144 | 2 | 75770002 | T | A | 0.14 | -0.289 | 3.87E-06 | AC005034.4 |
| rs10445937 | 2 | 60410521 | G | A | 0.40 | -0.213 | 4.18E-06 | MIR4432HG |
| rs12476132 | 2 | 60412769 | G | A | 0.40 | -0.213 | 4.18E-06 | MIR4432HG |
| rs12464462 | 2 | 60409281 | A | G | 0.40 | -0.212 | 4.63E-06 | MIR4432HG |
| rs10116133 | 9 | 7180759 | G | C | 0.22 | 0.248 | 4.66E-06 | AL161443.1 |
| rs243076 | 2 | 60390428 | G | A | 0.40 | -0.210 | 5.66E-06 | MIR4432HG |
| rs243068 | 2 | 60394240 | G | T | 0.40 | -0.210 | 5.66E-06 | MIR4432HG |
| rs243067 | 2 | 60394508 | G | C | 0.40 | -0.210 | 5.66E-06 | MIR4432HG |
| rs243066 | 2 | 60394631 | A | G | 0.40 | -0.210 | 5.66E-06 | MIR4432HG |
| rs243063 | 2 | 60395764 | A | T | 0.40 | -0.210 | 5.66E-06 | MIR4432HG |
| rs243062 | 2 | 60396541 | G | A | 0.40 | -0.210 | 5.66E-06 | MIR4432HG |
| rs243061 | 2 | 60396563 | T | C | 0.40 | -0.210 | 5.66E-06 | MIR4432HG |
| rs13011022 | 2 | 60399687 | G | A | 0.40 | -0.210 | 5.66E-06 | MIR4432HG |
| rs12988871 | 2 | 60403532 | G | A | 0.40 | -0.210 | 5.66E-06 | MIR4432HG |
| rs13009393 | 2 | 60406229 | G | A | 0.40 | -0.210 | 5.66E-06 | MIR4432HG |
| rs7585902 | 2 | 76731893 | A | G | 0.31 | -0.212 | 6.18E-06 | LRRTM4 |
| rs7584499 | 2 | 76734731 | T | G | 0.31 | -0.212 | 6.18E-06 | LRRTM4 |
| rs13406374 | 2 | 76734835 | G | A | 0.31 | -0.212 | 6.18E-06 | LRRTM4 |
| rs12713879 | 2 | 76738068 | T | A | 0.31 | -0.212 | 6.18E-06 | LRRTM4 |
| rs10181947 | 2 | 76738252 | C | T | 0.31 | -0.212 | 6.18E-06 | LRRTM4 |
| rs2540917 | 2 | 60381624 | T | C | 0.41 | -0.207 | 6.91E-06 | MIR4432HG |
| rs2540915 | 2 | 60382054 | C | T | 0.41 | -0.207 | 6.91E-06 | MIR4432HG |
| rs9967849 | 2 | 60382339 | C | T | 0.41 | -0.207 | 6.91E-06 | MIR4432HG |
| rs1553934 | 2 | 60382761 | G | C | 0.41 | -0.207 | 6.91E-06 | MIR4432HG |
| rs2540913 | 2 | 60383677 | T | C | 0.41 | -0.207 | 6.91E-06 | MIR4432HG |
| rs925483 | 2 | 60384150 | G | A | 0.41 | -0.207 | 6.91E-06 | MIR4432HG |
| rs925484 | 2 | 60384302 | C | A | 0.41 | -0.207 | 6.91E-06 | MIR4432HG |
| rs2137281 | 2 | 60384873 | C | T | 0.41 | -0.207 | 6.91E-06 | MIR4432HG |
| rs2137283 | 2 | 60385322 | C | A | 0.41 | -0.207 | 6.91E-06 | MIR4432HG |
| rs76453543 | 4 | 176813791 | T | C | 0.21 | 0.248 | 6.93E-06 | VEGFC |
| rs11154950 | 6 | 137971877 | A | G | 0.45 | -0.197 | 9.02E-06 | LINC02528 |
| rs11125842 | 2 | 60413672 | A | G | 0.42 | -0.202 | 9.04E-06 | MIR4432HG |
| rs969934 | 2 | 76741397 | C | A | 0.31 | -0.208 | 9.35E-06 | LRRTM4 |
| rs969933 | 2 | 76741469 | A | G | 0.31 | -0.208 | 9.35E-06 | LRRTM4 |
| rs12104736 | 2 | 60423581 | C | G | 0.42 | -0.202 | 9.57E-06 | MIR4432HG |
| rs12105503 | 2 | 60423607 | A | S | 0.42 | -0.202 | 9.57E-06 | MIR4432HG |
| rs55764830 | 2 | 60426130 | T | C | 0.42 | -0.202 | 9.57E-06 | MIR4432HG |
| rs1520484 | 3 | 46472191 | C | A | 0.28 | -0.214 | 9.92E-06 | LTF |
| rs243032 | 2 | 60376815 | G | A | 0.41 | -0.204 | 9.96E-06 | MIR4432HG |
| rs173373 | 2 | 60378784 | C | T | 0.41 | -0.204 | 9.96E-06 | MIR4432HG |
| rs2668734 | 2 | 60379174 | A | C | 0.41 | -0.204 | 9.96E-06 | MIR4432HG |
| rs243030 | 2 | 60379436 | C | T | 0.41 | -0.204 | 9.96E-06 | MIR4432HG |
| rs243029 | 2 | 60379519 | C | T | 0.41 | -0.204 | 9.96E-06 | MIR4432HG |
| rs13027161 | 2 | 60380593 | T | C | 0.41 | -0.204 | 9.96E-06 | MIR4432HG |

Abbreviations: SNP, single nucleotide polymorphism; p-tau181, phosphorylated tau at epitope 181; Chr, chromosome; Pos, position; Ref, reference allele; Alt, alternative allele; APOE, apolipoprotein E; MIR4432HG, MicroRNA 4432 Host Gene; LRRTM4, Leucine-Rich Repeat Transmembrane Neuronal 4; VEGFC, Vascular Endothelial Growth Factor C**; LINC02528, Long Intergenic Non-Protein Coding RNA 2528**; LTF, Lactotransferrin.

**Supplemental Figure 4.**  Linkage and GWAS signals on chromosome 1 for plasma p-tau181 levels. This figure shows the linkage analysis (black line) and GWAS (blue dots) results for plasma p-tau181 on chromosome 1. The red horizontal line marks the significant linkage threshold (LOD ≥ 3.3), where the strongest signal appears at rs7365459 (LOD = 3.53). The dashed blue line indicates the GWAS suggestive threshold (*p* = 1 × 10⁻⁵). Notably, the top GWAS hit is rs61798463 in this region with a *p*-value of 1.25 × 10^−4^. Abbreviations: p-tau181, phosphorylated tau at epitope 181; GWAS, genome-wide association study; LOD, logarithm of the odds.

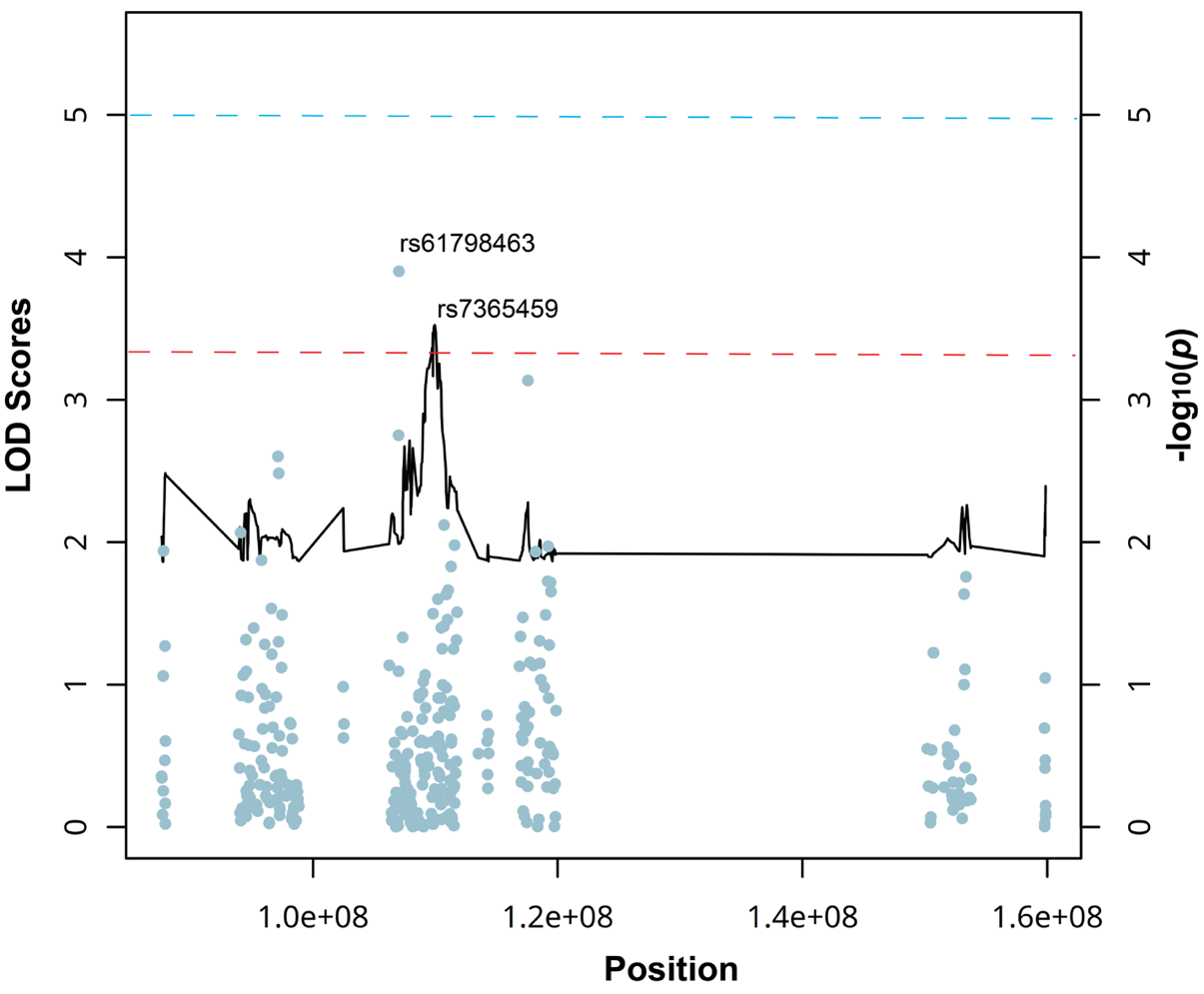

**Supplemental Figure 5.** Linkage analysis of plasma p-tau181 levels in the Amish using 21-bit (blue) and 25-bit pedigrees (red). The blue horizontal line represents the suggestive linkage threshold at LOD scores ≥ 1.86, while the red horizontal line marks the significant linkage threshold at LOD scores ≥ 3.3. Abbreviations: p-tau181, phosphorylated tau at epitope 181; LOD, logarithm of the odds; cM, centimorgan.

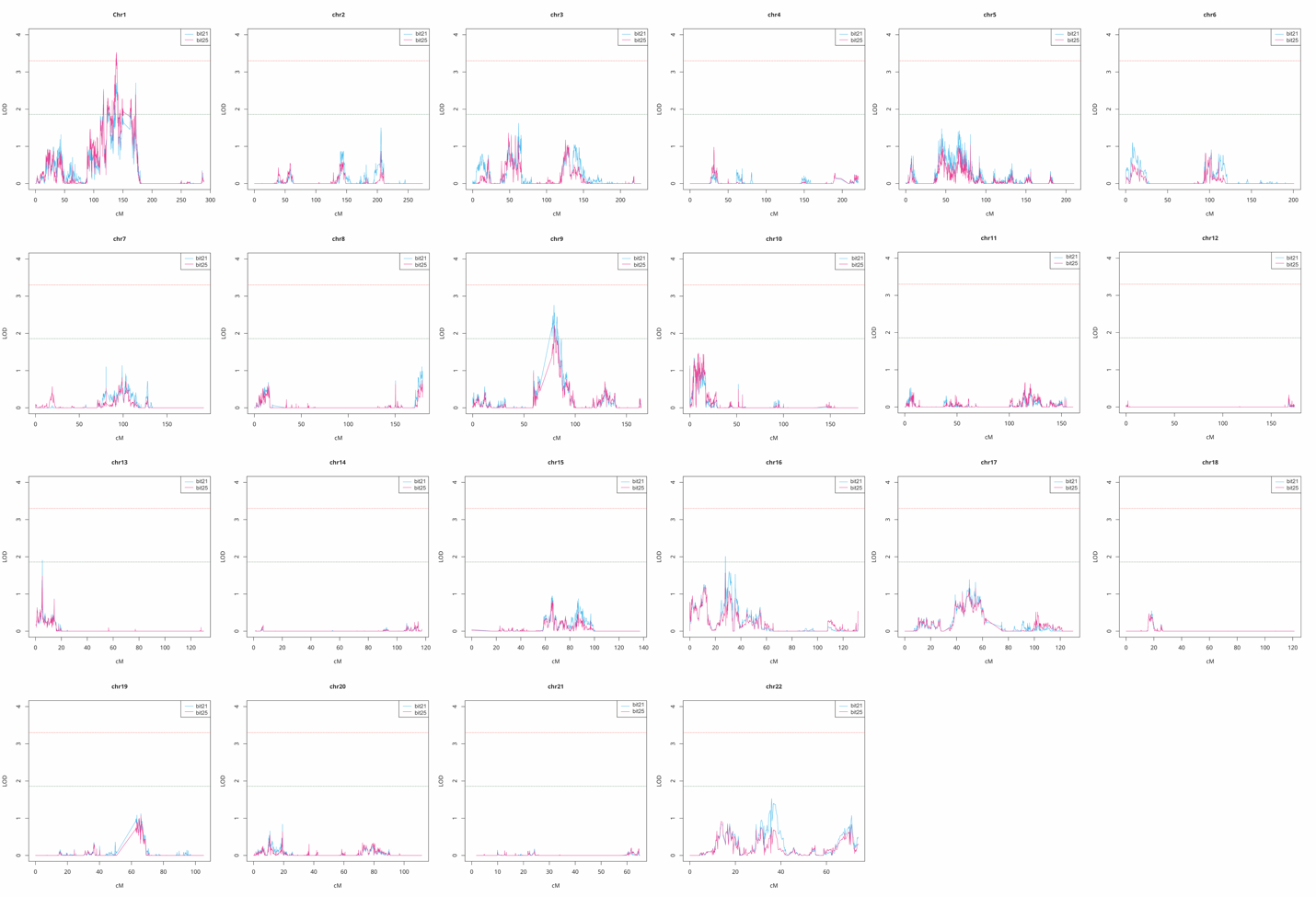

**Supplemental Figure 6.** Manhattan plot (A) and quantile-quantile plot (B) of plasma p-tau181/Aβ42 ratio in the Amish. In the Manhattan plot, the blue line marks the genome-wide suggestive threshold (*p* = 1 × 10⁻⁵), while the red line represents the genome-wide significance threshold (*p* = 5 × 10⁻⁸). Abbreviations: Aβ, amyloid beta; p-tau181, phosphorylated tau at epitope 181; LINC01378, Long Intergenic Non‑Protein Coding RNA 1378; **HLA-DQB1**, Human Leukocyte Antigen DQ Beta 1; **TRAPPC9**, Trafficking Protein Particle Complex 9; **GRHPR**, Glyoxylate Reductase/Hydroxypyruvate Reductase; **POU2F3**, POU Class 2 Homeobox 3; **NTN1**, Netrin 1; APOE, apolipoprotein E.

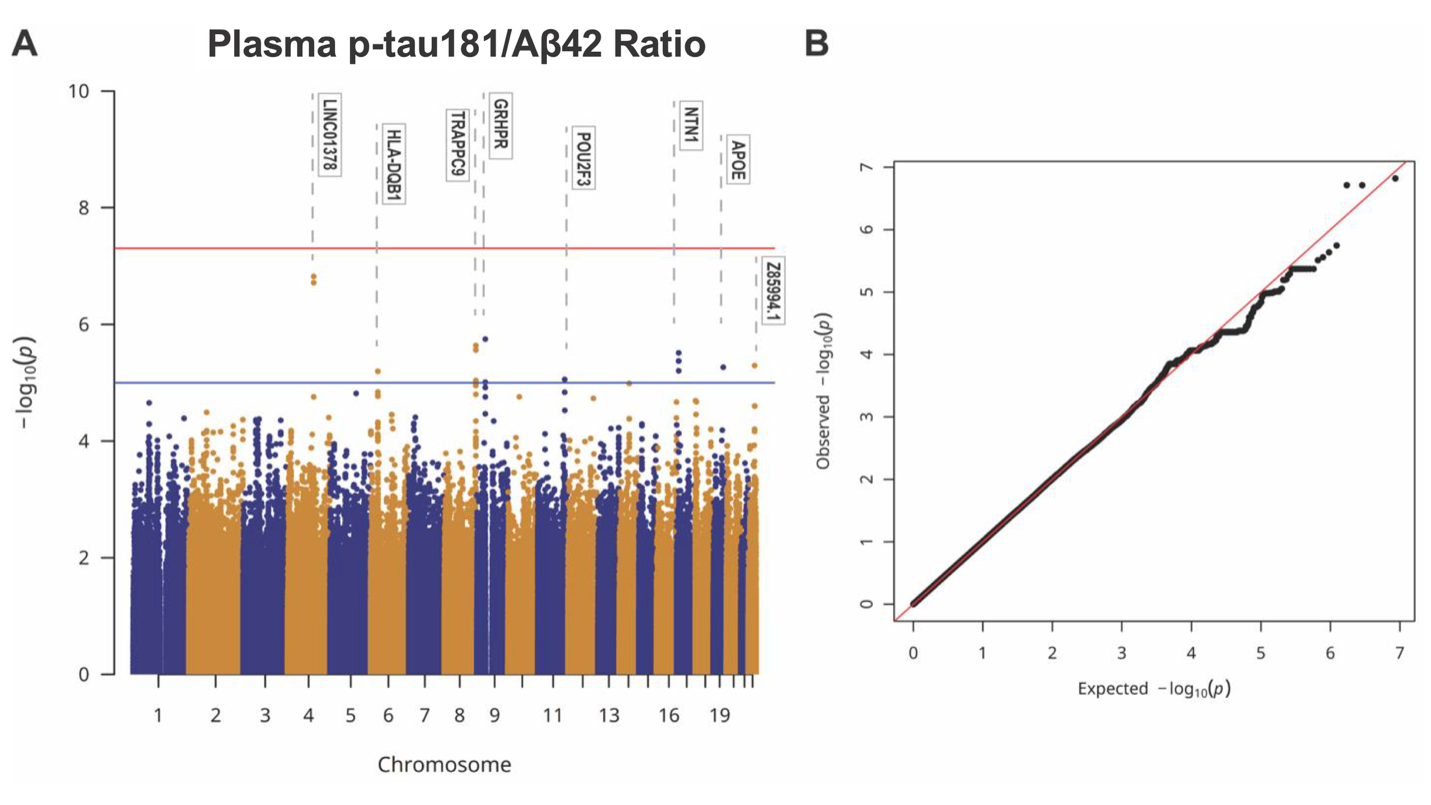

**Supplemental Table 3**. SNPs associated with the plasma p-tau181/Aβ42 ratio in the Amish (*p* ≤ 1 × 10⁻⁵).

| **Variant** | **Chr** | **Pos** | **Ref** | **Alt** | **MAF** | **Estimate** | ***p*** | **Overlapped/**  **Nearby Gene** |
| --- | --- | --- | --- | --- | --- | --- | --- | --- |
| rs200118070 | 4 | 117411035 | - | T | 0.20 | -0.351 | 1.51E-07 | LINC01378 |
| rs17861871 | 4 | 117407354 | A | T | 0.20 | -0.348 | 1.94E-07 | LINC01378 |
| rs17861878 | 4 | 117409299 | C | G | 0.20 | -0.348 | 1.94E-07 | LINC01378 |
| rs7866763 | 9 | 37411382 | C | A | 0.10 | -0.427 | 1.79E-06 | GRHPR |
| rs7840390 | 8 | 140354029 | G | T | 0.30 | -0.276 | 2.31E-06 | TRAPPC9 |
| rs7000128 | 8 | 140443202 | A | C | 0.33 | -0.266 | 2.76E-06 | TRAPPC9 |
| rs774957404 | 17 | 9100090 | T | - | 0.24 | -0.299 | 3.09E-06 | NTN1 |
| rs9899718 | 17 | 9098950 | T | C | 0.24 | -0.293 | 4.25E-06 | NTN1 |
| rs9894396 | 17 | 9099513 | C | T | 0.24 | -0.293 | 4.25E-06 | NTN1 |
| rs9892896 | 17 | 9099602 | G | A | 0.24 | -0.293 | 4.25E-06 | NTN1 |
| rs9902695 | 17 | 9100384 | A | G | 0.24 | -0.293 | 4.25E-06 | NTN1 |
| rs9915510 | 17 | 9101024 | T | C | 0.24 | -0.293 | 4.25E-06 | NTN1 |
| rs57602873 | 17 | 9102578 | G | A | 0.24 | -0.293 | 4.25E-06 | NTN1 |
| rs9905655 | 17 | 9104176 | A | G | 0.24 | -0.293 | 4.25E-06 | NTN1 |
| rs9912864 | 17 | 9105233 | C | T | 0.24 | -0.293 | 4.25E-06 | NTN1 |
| rs9906185 | 17 | 9105898 | C | T | 0.24 | -0.293 | 4.25E-06 | NTN1 |
| rs6007197 | 22 | 44369771 | G | A | 0.42 | 0.240 | 5.10E-06 | Z85994.1 |
| rs429358 | 19 | 44908684 | T | C | 0.14 | 0.360 | 5.44E-06 | APOE |
| rs871827 | 17 | 9090584 | A | C | 0.24 | -0.288 | 6.26E-06 | NTN1 |
| rs7744020 | 6 | 32658353 | G | A | 0.31 | -0.253 | 6.39E-06 | HLA-DQB1 |
| rs35779483 | 6 | 32658925 | A | G | 0.31 | -0.253 | 6.39E-06 | HLA-DQB1 |
| rs61898318 | 11 | 120300662 | G | A | 0.10 | -0.376 | 8.82E-06 | POU2F3 |
| rs12155853 | 8 | 140360979 | C | T | 0.32 | -0.255 | 9.16E-06 | TRAPPC9 |
| rs13265382 | 8 | 140379063 | T | C | 0.30 | -0.257 | 9.75E-06 | TRAPPC9 |
| rs10875452 | 8 | 140388117 | G | A | 0.30 | -0.257 | 9.75E-06 | TRAPPC9 |
| rs28410628 | 8 | 140403707 | C | A | 0.30 | -0.257 | 9.75E-06 | TRAPPC9 |
| rs7876059 | 9 | 37394153 | T | A | 0.10 | -0.388 | 9.84E-06 | LINC01627 |
| rs7860922 | 9 | 37394376 | G | C | 0.10 | -0.388 | 9.84E-06 | LINC01627 |

Abbreviations: SNP, single nucleotide polymorphism; Aβ, amyloid beta; p-tau181, phosphorylated tau at epitope 181; Chr, chromosome; Pos, position; Ref, reference allele; Alt, alternative allele; LINC01378, Long Intergenic Non‑Protein Coding RNA 1378; **GRHPR**, Glyoxylate Reductase/Hydroxypyruvate Reductase; **TRAPPC9**, Trafficking Protein Particle Complex 9; **NTN1**, Netrin 1; APOE, apolipoprotein E; **HLA-DQB1**, Human Leukocyte Antigen DQ Beta 1; **POU2F3**, POU Class 2 Homeobox 3; LINC01627, Long Intergenic Non‑Protein Coding RNA 1627.

**Supplemental Figure 7.** Linkage analysis of plasma p-tau181/Aβ42 ratio using 25-bit pedigrees in the Amish identified one significant locus (LOD = 1.91) on chromosome 4. The blue horizontal line represents the suggestive linkage threshold at LOD scores ≥ 1.86. Abbreviations: Aβ, amyloid beta; p-tau181, phosphorylated tau at epitope 181; LOD, logarithm of the odds; cM, centimorgan.

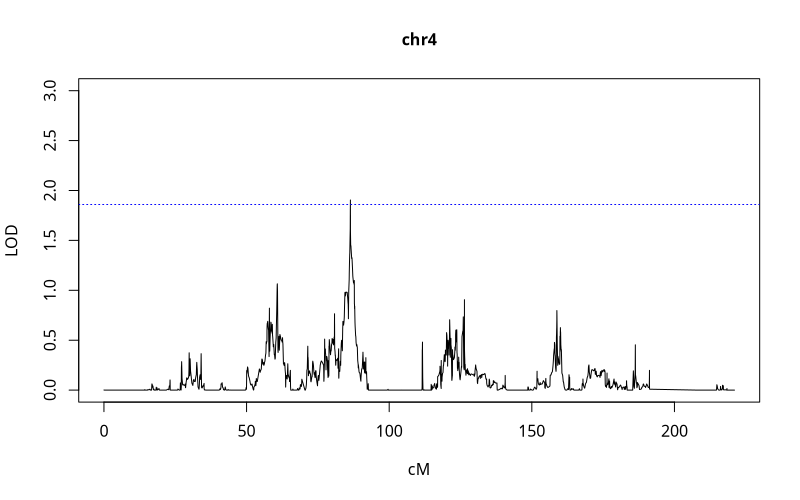

**Supplemental Figure 8.** Linkage analysis of the plasma p-tau181/Aβ42 ratio in the Amish using 25-bit pedigrees. The blue horizontal line represents the suggestive linkage threshold at LOD scores ≥ 1.86. Abbreviations: Aβ, amyloid beta; p-tau181, phosphorylated tau at epitope 181; LOD, logarithm of the odds; cM, centimorgan.

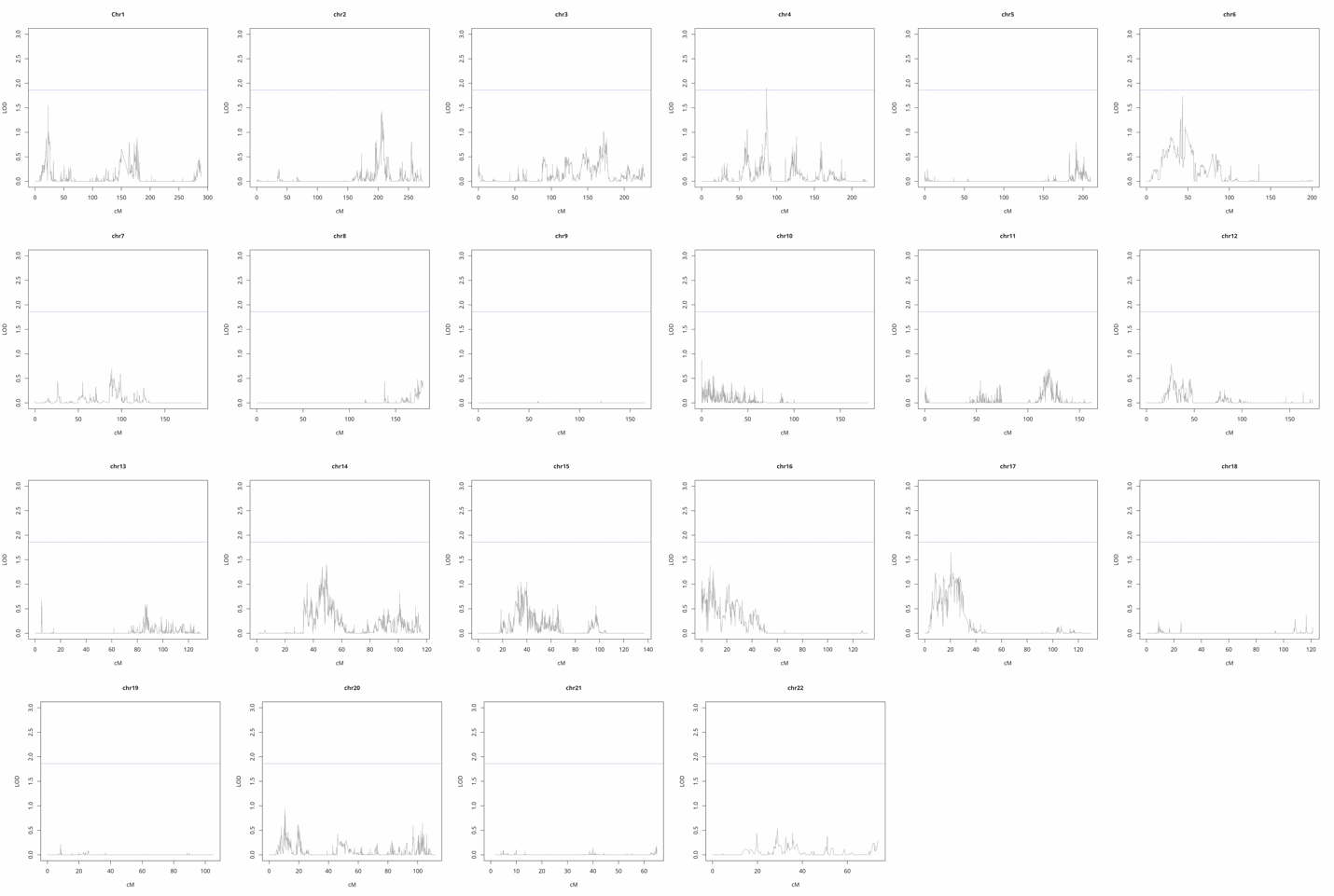

**Supplemental Figure 9.** Manhattan plot (A) and quantile-quantile plot (B) of plasma Aβ42 in the Amish. In the Manhattan plot, the blue line marks the genome-wide suggestive threshold (*p* = 1 × 10⁻⁵), while the red line represents the genome-wide significance threshold (*p* = 5 × 10⁻⁸). Abbreviations: Aβ, amyloid beta; CCNT2P, cyclin T2 pseudogene1; CERS6; Ceramide Synthase 6; LINC02653, long intergenic non-protein coding RNA 2653; PPHLN1, periphilin-1; RAB4B, member RAS oncogene family; NCAM2, neural cell adhesion molecule 2.

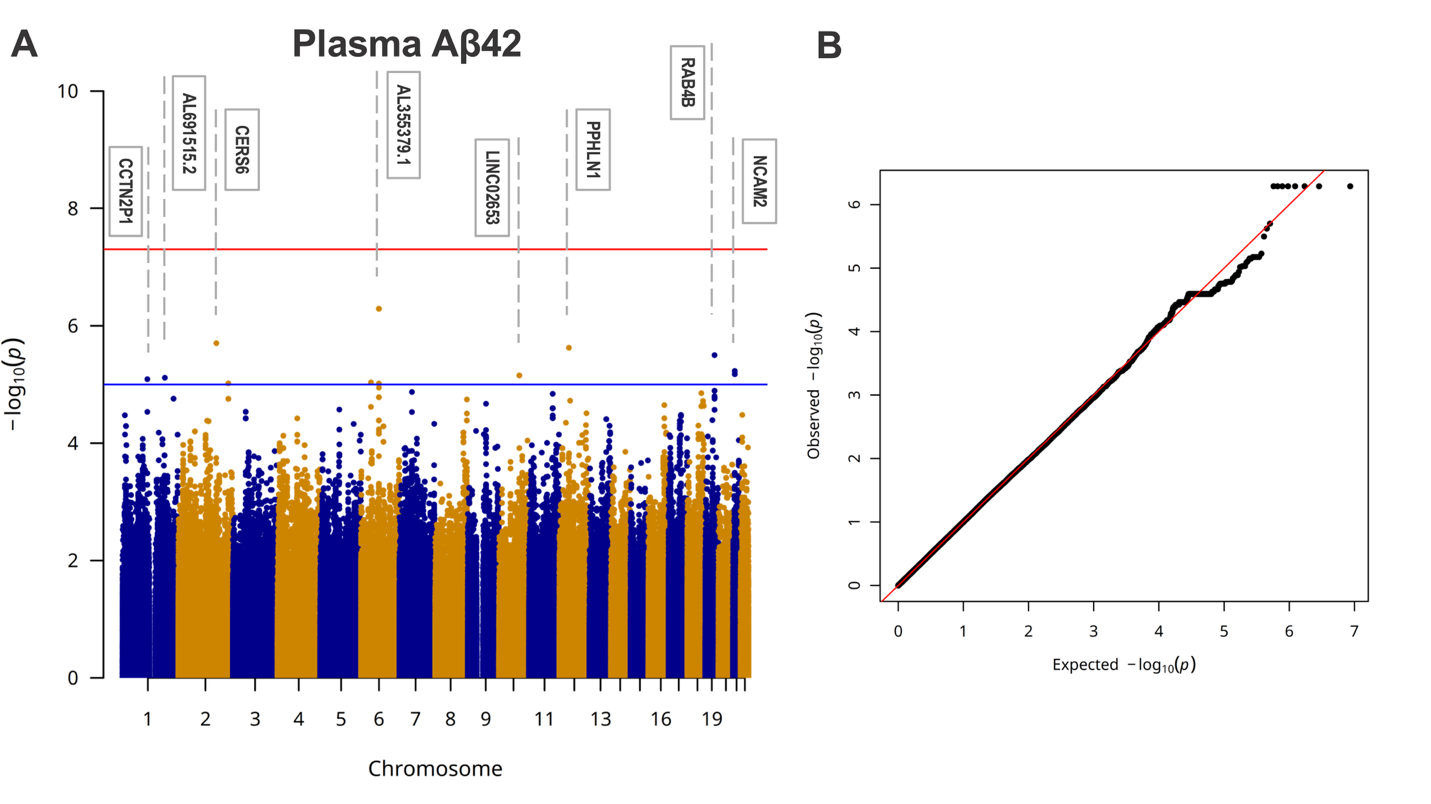

**Supplemental Table 4**. SNPs associated with plasma Aβ42 in the Amish (*p* ≤ 1 × 10⁻⁵).

| **Variant** | **Chr** | **Pos** | **Ref** | **Alt** | **MAF** | **Estimate** | ***p*** | **Overlapped/**  **Nearby Gene** |
| --- | --- | --- | --- | --- | --- | --- | --- | --- |
| rs72895663 | 6 | 79339973 | A | G | 0.25 | 0.263 | 5.14E-07 | AL355379.1 |
| rs11753238 | 6 | 79341988 | G | T | 0.25 | 0.263 | 5.14E-07 | AL078601.1 |
| rs72895666 | 6 | 79342047 | A | G | 0.25 | 0.263 | 5.14E-07 | AL078601.1 |
| rs72895671 | 6 | 79343354 | G | T | 0.25 | 0.263 | 5.14E-07 | AL078601.1 |
| rs72895672 | 6 | 79343933 | C | T | 0.25 | 0.263 | 5.14E-07 | AL078601.1 |
| rs72895676 | 6 | 79346657 | G | A | 0.25 | 0.263 | 5.14E-07 | AL078601.1 |
| rs11757083 | 6 | 79347633 | C | T | 0.25 | 0.263 | 5.14E-07 | AL078601.1 |
| rs140005740 | 6 | 79352306 | T | C | 0.25 | 0.263 | 5.14E-07 | AL078601.1 |
| rs17251018 | 2 | 168731685 | G | A | 0.45 | -0.216 | 1.99E-06 | CERS6 |
| rs7978993 | 12 | 42286849 | G | A | 0.28 | -0.241 | 2.38E-06 | PPHLN1 |
| rs78469339 | 19 | 40794564 | A | T | 0.15 | 0.297 | 3.18E-06 | RAB4B |
| rs2826860 | 21 | 21487004 | G | A | 0.20 | -0.257 | 5.91E-06 | NCAM2 |
| rs233755 | 21 | 21481832 | G | A | 0.20 | -0.255 | 6.68E-06 | NCAM2 |
| rs2009029 | 21 | 21483571 | G | A | 0.20 | -0.255 | 6.68E-06 | NCAM2 |
| rs114308 | 21 | 21484961 | G | T | 0.20 | -0.255 | 6.68E-06 | NCAM2 |
| rs233761 | 21 | 21485008 | C | T | 0.20 | -0.255 | 6.68E-06 | NCAM2 |
| rs1773395 | 10 | 90523232 | C | A | 0.29 | -0.222 | 7.05E-06 | LINC02653 |
| rs10458742 | 10 | 90539802 | G | C | 0.29 | -0.222 | 7.05E-06 | LINC02653 |
| rs4598489 | 1 | 188790814 | C | T | 0.37 | -0.209 | 7.72E-06 | AL691515.2 |
| rs7515000 | 1 | 111001126 | C | T | 0.19 | 0.257 | 8.16E-06 | CCNT2P1 |
| rs833810 | 6 | 44066954 | C | A | 0.36 | -0.214 | 9.30E-06 | SCIRT |
| rs9381285 | 6 | 44067629 | G | A | 0.36 | -0.214 | 9.30E-06 | SCIRT |
| rs833812 | 6 | 44067844 | G | A | 0.36 | -0.214 | 9.30E-06 | SCIRT |
| rs60630276 | 2 | 221934143 | G | C | 0.32 | 0.213 | 9.60E-06 | AC011752.1 |
| rs7770603 | 6 | 79326698 | G | A | 0.17 | 0.271 | 9.65E-06 | AL355379.1 |

Abbreviations: SNP, single nucleotide polymorphism; Aβ, amyloid beta; Chr, chromosome; Pos, position; Ref, reference allele; Alt, alternative allele; CERS6; Ceramide Synthase 6; PPHLN1, periphilin-1; RAB4B, member RAS oncogene family; NCAM2, neural cell adhesion molecule 2; LINC02653, long intergenic non-protein coding RNA 2653; CCNT2P, cyclin T2 pseudogene1; SCIRT, stem cell inhibitory RNA transcript.
